# Humoral responses against HDL particles are linked to lipoprotein traits, atherosclerosis occurrence, inflammation and pathogenic pathways during the earliest stages of arthritis

**DOI:** 10.1101/2022.08.12.22278696

**Authors:** Javier Rodríguez-Carrio, Mercedes Alperi-López, Patricia López, Ángel I. Pérez-Álvarez, George A. Robinson, Sara Alonso-Castro, Núria Amigó, Fabiola Atzeni, Ana Suárez

## Abstract

**Objective:** chronic inflammation and immune dysregulation are crucial mechanisms for atherosclerosis in rheumatoid arthritis (RA). Recent evidence suggests a link via humoral responses against high-density lipoproteins (HDL). However, their specificity, clinical relevance and emergence along disease course are unknown, especially during the earliest phases of RA.

**Methods:** IgG and IgM serum levels of antibodies against HDL (anti-HDL) and Apolipoprotein A1 (anti-ApoA1) were measured in 82 early RA patients, 14 arthralgia individuals and 96 controls. Established RA patients (n=42) were included for validation. Atherosclerosis and vascular stiffness were measured by Doppler-ultrasound. Lipoprotein content, particle numbers and size were measured by H-NMR. Cytokines were measured by immunoassays. A cardiometabolic-related protein panel was evaluated using high-throughput targeted proteomics.

**Results:** anti-HDL and anti-ApoA1 responses were increased in early RA compared to controls (both p<0.001) and were comparable to established disease. Only anti-ApoA1 antibodies were increased in arthralgia. IgG anti-HDL and anti-ApoA1 were associated with unfavourable lipoprotein traits in RA and arthralgia, respectively. A similar picture was observed for inflammatory mediators. No associations with clinical features or risk factors were found. IgG anti-HDL were independently associated with atherosclerosis occurrence in early RA, and outperformed patient stratification over conventional algorithms (mSCORE) and their anti-ApoA1 counterparts. Anti-HDL antibodies correlated with proteins involved in immune activation, remodelling, and lipid metabolism pathways in early RA.

**Conclusion:** humoral responses against HDL particles are an early event along arthritis course, although quantitative and qualitative differences can be noticed among stages. These differences informed distinct capacities as biomarkers and underlying pathogenic circuits.

## INTRODUCTION

Rheumatoid Arthritis (RA) has been consistently associated with an increased cardiovascular disease (CVD) occurrence compared to the general population, due to an accelerated development and progression of atherosclerosis (1). This risk excess cannot be fully explained by traditional CV risk factors alone, thus pointing to the involvement of non- traditional CV risk factors (2). However, these are poorly characterized until date, which limits CV risk stratification and represents an urgent clinical need.

Compelling evidence has demonstrated a protective effect of high-density lipoprotein- cholesterol (HDL-C) levels on CVD in the general population, although the picture in RA seems to be more complex (3). Low HDL-C levels were initially considered as a traditional risk factor, due to its impaired ability to remove cholesterol excess, although recent evidence has challenged this notion. A number of non-canonical functions, such as anti-oxidant, anti- inflammatory, anti-apoptotic and anti-thrombotic properties have been reported to contribute to its anti-atherogenic effect (4). Inflammation is known to cause both changes in the lipoprotein levels as well as in their protein composition and non-canonical functions (5–7). Furthermore, different immunosuppressive agents are known to modulate lipoprotein levels and functions to variable, different degrees (5,8), thus emphasizing the active involvement of specific immune pathways. Therefore, HDL-C levels do not necessarily correlate with protective functions, especially during inflammation, leading to the concept of HDL dysfunction (9). However, important gaps remain in the understanding on the crosstalk between HDL and inflammation and immune pathways, especially beyond HDL-C levels.

A potential role of the humoral response in this setting has emerged in recent years. The presence of IgG antibodies against HDL (anti-HDL) and its components has been demonstrated by our group (10–13) and others (14–17) in several inflammatory conditions. We have found that the IgG anti-HDL response is increased in RA patients with established disease, linked to inflammatory burden and CVD history (12). However, whether these antibodies are present at disease onset or are a consequence of the disease course and/or changes in HDL due to CVD occurrence remains unknown. This is of pivotal relevance to evaluate their potential capacity for improving risk stratification, especially during the early stages. Importantly, autoimmune responses are known to predate disease onset in RA (18,19). Moreover, since HDL are complex structures that need to be studied beyond HDL- C levels, there is a need for multifaceted approaches that include HDL composition, size, functionality, and underlying pathogenic circuits to better understand the relevance of IgG anti-HDL responses, especially from a non-traditional perspective. Finally, although the analysis of anti-ApoA1 responses has become popular, evidence from lupus patients suggests that anti-HDL and anti-ApoA1 may not be used interchangeably (14,15). However, head-to- head comparative analyses are much awaited.

Taken together, we hypothesize that IgG anti-HDL can be considered a non-traditional risk factor in early RA, which can inform the lipoprotein-inflammation crosstalk and account for the HDL dysfunction phenomenon, hence providing added value for improving risk stratification. Then, the main aims of this study were (i) to characterize the humoral response against HDL structure during the early phases of RA, (ii) to evaluate the associations between the humoral response against HDL and lipoprotein features, including size, content, and functionality, (iii) to evaluate their potential role as a biomarker for risk stratification, and (iv) to characterize the underlying pathogenic circuits by a proteomic analysis.

## MATERIAL AND METHODS

### Study participants

Our study involved 82 early RA patients (2010 ACR/EULAR criteria), 14 arthralgia individuals and 96 age- and sex-matched individuals as healthy controls (HC). Subclinical atherosclerosis and vascular stiffness were measured by Doppler ultrasound assessments. Detailed information about recruitment and clinical procedures can be found in the supplementary material (Supplementary material and methods).

### Quantification of antibodies against HDL particles

Levels of antibodies against HDL and ApoA1 (both IgG and IgM isotypes) were quantified in serum samples as previously described (12) with slight modifications (Supplementary material and methods). Antibody levels were expressed as arbitrary units (AU).

### Assessment of PON1 activity

PON1 activity in serum was quantified by means of an enzymatic assay according to Eckerson et al. with slight modifications as reported by our group (10) (Supplementary material and methods).

### Lipoprotein characterization

An advanced lipoprotein characterization by means of the H-NMR-based Liposcale test was performed (Supplementary material and methods).

### Proteomic analysis

Levels of 92 proteins involved in cardiovascular disease were evaluated in serum by a proteomic approach using the Proximity Extension Assay (PEA) Olink technology (Supplementary material and methods).

### Statistical analyses

Continuous variables were expressed as median (interquartile range) or mean±standard deviation, whereas categorical ones were expressed as n(%). Differences among groups were evaluated by one-way ANOVA, Mann-Withney U, Kruskal-Wallis or χ2 tests, as appropriate. Statistical analyses were carried out under SPSS v. 27 and R v.4.1.3. Detailed information on statistical analysis can be found in the supplementary material (Supplementary material and methods).

## RESULTS

### Anti-HDL and anti-ApoA1 humoral responses emerge during the earliest stages of RA

The levels of anti-HDL and anti-ApoA1 antibodies (both IgG and IgM isotypes) were measured in serum samples from 82 early RA patients, 14 CSA individuals and 96 HC (Supplementary Table 1).

IgG and IgM anti-HDL antibodies were found to be increased in RA patients compared to HC, and similar findings were observed for anti-ApoA1 responses (Figure 1A). RA patients also exhibited higher IgG anti-HDL levels compared to CSA individuals. Higher IgG anti- ApoA1 levels were observed in the CSA group compared to HC (Figure 1A), and levels of IgG anti-HDL were also numerically higher in this group compared to HC (298.89 (416.16) vs 180.09 (564.30) AU). When RA patients were compared to the validation cohort of long-lasting, established RA patients (LRA) (Supplementary Table 2), no differences were found in any of the antibodies studied (Supplementary Figure 1). Treatments did not influence antibody levels in this cohort (all p>0.050). No correlations between each antibody and the corresponding total Ig serum levels (IgG or IgM) were retrieved in any group, and between-group differences remained after correcting by total Ig levels.

**Figure 1:**
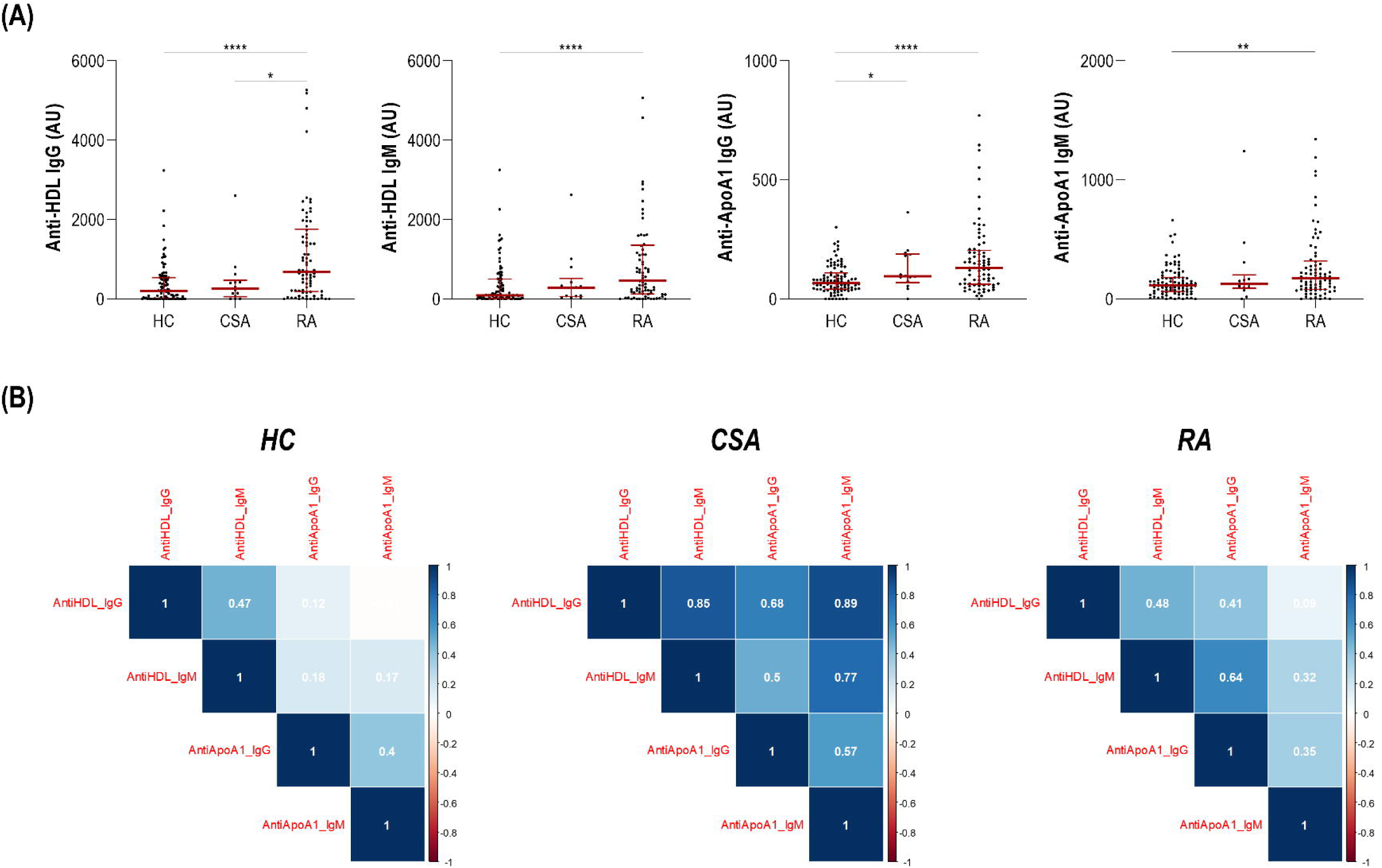
Levels of antibodies against HDL particles across study groups. (A) Levels of IgG anti-HDL and anti-ApoA1 (both IgG and IgM) in HC, CSA individuals and early RA patients are shown. Bars represent 25^th^ percentile (lower), median and 75^th^ percentile (upper). Differences were assessed by Kruskal-Wallis tests with Dunn-Bonferroni post-hoc tests. The p-values from the latter were indicated as follows: * p<0.050, ** p<0.010 and *** p<0.001. (B) The associations among different antibodies (isotypes and/or specificities) were studied across study groups in correlograms. Correlation coefficients for each pair of variables are shown (white). Colour gradient varied blue (positive correlations) to red (negative correlations).

Next, the associations between levels of antibodies were studied. The CSA group showed higher correlations between specificities (IgG anti-HDL vs IgG anti-ApoA1), whereas these correlations were of a much lower degree in the RA group (Figure 1B). An equivalent picture was found between isotypes from the same specificity.

These results confirm that humoral responses against HDL particles are present already during the earliest phases of RA, and no differences between early and established RA were found. On the contrary, the CSA groups was hallmarked by a heterogeneous profile of humoral responses, with differences in its extent and specificities compared to clinical disease.

### Anti-HDL and anti-ApoA1 antibodies exhibit distinct associations with lipoprotein profiles and inflammatory mediators in CSA and RA

Next, the associations between antibodies against HDL particles and lipoprotein profiles (Supplementary Table 3) obtained by H-NMR were analysed. No major differences in lipoprotein assessments were found across groups. IgG anti-HDL levels were correlated with lipoprotein content in very low-, intermediate- and high-density lipoproteins, as well as negatively with HDL particle number in RA patients (Table 1). Of note, these associations were mostly attributed to the HDL small particle subclass, which was strongly positively correlated with PON1 activity in this group (Supplementary Figure 3) (Supplementary Table 4). No associations with IgM isotype or anti-ApoA1 antibodies were registered. Although no associations with IgG anti-HDL were found in CSA individuals, IgG anti-ApoA1 levels negatively paralleled HDL content, particle number and size distribution in CSA individuals (Table 1), thus mirroring those of the IgG anti-HDL in the RA group. No associations were registered in HC.

**Table 1:** Associations between antibodies against HDL and lipoprotein features. Associations between levels of antibodies against HDL or ApoA1 and lipoprotein features (particle content, particle size and subclasses) were analysed by Spearman’s rank tests in CSA and RA groups. Correlation coefficients (r) and p-values are shown. Those reaching statistical significance are highlighted in bold.

|  | CSA |  |  |  | RA |  |  |  |
| --- | --- | --- | --- | --- | --- | --- | --- | --- |
|  | Anti-HDL<br>IgG | Anti-HDL<br>IgM | Anti-ApoA1<br>IgG | Anti-ApoA1<br>IgM | Anti-HDL<br>IgG | Anti-HDL<br>IgM | Anti-ApoA1<br>IgG | Anti-ApoA1<br>IgM |
| *Particle content* |  |  |  |  |  |  |  |  |
| VLDL-C | r=-0.011<br>p=0.970 | r=-0.011<br>p=0.970 | r=0.371<br>p=0.191 | r=-0.407<br>p=0.149 | **r=0.273**<br>**p=0.013** | r=0.125.<br>p=0.262 | r=0.012<br>p=0.913 | r=-0.044<br>p=0.697 |
| IDL-C | r=-0.018<br>p=0.652 | r=-0.191<br>p=0.513 | r=0.349<br>p=0.221 | r=0.015<br>p=0.958 | **r=0.300**<br>**p=0.006** | r=0.190<br>p=0.088 | r=0.096<br>p=0.391 | r=0.064<br>p=0.566 |
| LDL-C | r=-0.029<br>p=0.923 | r=0.213<br>p=0.464 | r=0.345<br>p=0.215 | r=0.385<br>p=0.175 | r=-0.090<br>p=0.423 | r=0.065<br>p=0.536 | r=0.029<br>p=0.794 | r=0.071<br>p=0.528 |
| HDL-C | r=-0.136<br>p=0.642 | r=-0.138<br>p=0.637 | **r=-0.411**<br>**p=0.040** | r=-0.113<br>p=0.702 | **r=-0.302**<br>**p=0.006** | r=-0.127<br>p=0.256 | r=0.059<br>p=0.596 | r=0.102<br>p=0.362 |
| VLDL-TG | r=-0.015<br>p=0.958 | r=-0.200<br>p=0.493 | r=0.284<br>p=0.326 | r=-0.477<br>p=0.085 | r=0.177<br>p=0.112 | r=0.049<br>p=0.664 | r=-0.103<br>p=0.357 | r=-0.076<br>p=0.499 |
| IDL- TG | r=-0.055<br>p=0.852 | r=-0.244<br>p=0.401 | r=0.231<br>p=0.427 | r=0.002<br>p=0.992 | **r=0.226**<br>**p=0.041** | r=0.161<br>p=0.150 | r=0.057<br>p=0.611 | r=0.047<br>p=0.678 |
| LDL- TG | r=0.079<br>p=0.788 | r=-0.086<br>p=0.771 | r=0.455<br>p=0.102 | r=0.270<br>p=0.350 | **r=0.210**<br>**p=0.058** | r=0.192<br>p=0.084 | r=0.218<br>p=0.049 | r=0.151<br>p=0.176 |
| HDL- TG | r=-0.084<br>p=0.776 | r=-0.446<br>p=0.110 | r=0.200<br>p=0.493 | r=-0.178<br>p=0.543 | r=0.099<br>p=0.376 | r=0.149<br>p=0.180 | r=0.095<br>p=0.397 | r=0.125<br>p=0.265 |
| **Particle number** |  |  |  |  |  |  |  |  |
| VLDL-P<br>(nmol/l) | r=-0.013<br>p=0.964 | r=-0.160<br>p=0.584 | r=0.332<br>p=0.246 | r=-0.486<br>p=0.078 | r=0.197<br>p=0.075 | r=0.077<br>p=0.464 | r=-0.075<br>p=0.506 | r=-0.060<br>p=0.593 |
| Large | r=-0.059<br>p=0.840 | r=-0.178<br>p=0.543 | r=0.253<br>p=0.383 | r=-0.516<br>p=0.059 | r=0.169<br>p=0.130 | r=0.020<br>p=0.859 | r=-0.108<br>p=0.333 | r=-0.117<br>p=0.294 |
| Medium | r=0.040<br>p=0.893 | r=-0.042<br>p=0.887 | r=0.459<br>p=0.098 | r=-0.437<br>p=0.118 | r=0.246<br>p=0.026 | r=0.031<br>p=0.779 | r=-0.051<br>p=0.647 | r=-0.061<br>p=0.588 |
| Small | r=-0.048<br>p=0.869 | r=-0.187<br>p=0.523 | r=0.266<br>p=0.358 | r=-0.486<br>p=0.078 | r=0.192<br>p=0.085 | r=0.078<br>p=0.484 | r=-0.071<br>p=0.525 | r=-0.054<br>p=0.627 |
| LDL-P<br>(nmol/l) | r=-0.031<br>p=0.917 | r=0.196<br>p=0.503 | r=0.327<br>p=0.253 | r=0.275<br>p=0.342 | r=-0.072<br>p=0.521 | r=0.070<br>p=0.531 | r=0.027<br>p=0.807 | r=0.071<br>p=0.528 |
| Large | r=0.165<br>p=0.573 | r=0.156<br>p=0.594 | r=0.415<br>p=0.140 | r=0.418<br>p=0.137 | r=-0.024<br>p=0.829 | r=0.152<br>p=0.172 | r=0.166<br>p=0.137 | r=0.200<br>p=0.071 |
| Medium | r=0.077<br>p=0.794 | r=0.143<br>p=0.626 | r=0.415<br>p=0.141 | r=0.552<br>p=0.041 | r=-0.024<br>p=0.829 | r=0.136<br>p=0.221 | r=0.165<br>p=0.139 | r=0.167<br>p=0.133 |
| Small | r=-0.022<br>p=0.940 | r=0.187<br>p=0.523 | r=0.341<br>p=0.233 | r=0.086<br>p=0.771 | r=-0.127<br>p=0.254 | r=0.041<br>p=0.716 | r=-0.130<br>p=0.246 | r=0.011<br>p=0.920 |
| HDL-P<br>(mmol/l) | r=-0.180<br>p=0.537 | r=-0.275<br>p=0.342 | **r=-0.584**<br>**p=0.028** | r=-0.239<br>p=0.410 | **r=-0.356**<br>**p=0.001** | r=-0.206<br>p=0.064 | r=0.029<br>p=0.796 | r=0.081<br>p=0.469 |
| Large | r=-0.158<br>p=0.589 | r=0.002<br>p=0.994 | r=0.130<br>p=0.659 | r=0.301<br>p=0.296 | r=-0.008<br>p=0.943 | r=0.134<br>p=0.232 | r=0.214<br>p=0.054 | **r=0.243**<br>**p=0.029** |
| Medium | r=-0.139<br>p=0.637 | r=-0.081<br>p=0.782 | r=-0.270<br>p=0.350 | r=0.288<br>p=0.318 | r=-0.139<br>p=0.213 | r=0.008<br>p=0.944 | r=0.209<br>p=0.084 | r=0.215<br>p=0.054 |
| Small | r=-0.202<br>p=0.488 | r=-0.327<br>p=0.253 | **r=-0.581**<br>**p=0.021** | r=0.138<br>p=0.637 | **r=-0.388**<br>**p<0.001** | **r=-0.290**<br>**p=0.008** | r=-0.050<br>p=0.658 | r=-0.008<br>p=0.940 |

| *Particle diameter (nm)* |  |  |  |  |  |  |  |  |
| --- | --- | --- | --- | --- | --- | --- | --- | --- |
| VLDL | r=0.205<br>p=0.483 | r=0.187<br>p=0.523 | r=-0.086<br>p=0.771 | r=0.204<br>p=0.483 | r=0.057<br>p=0.609 | r=0.156<br>p=0.161 | r=0.055<br>p=0.624 | r=-0.037<br>p=0.0740 |
| LDL | r=-0.004<br>p=0.988 | r=0.107<br>p=0.714 | r=0.051<br>p=0.864 | r=0.389<br>p=0.169 | r=0.135<br>p=0.228 | r=0.158<br>p=0.157 | **r=0.299**<br>**p=0.006** | **r=0.253**<br>**p=0.022** |
| HDL | r=0.427<br>p=0.127 | r=0.525<br>p=0.054 | r=0.455<br>p=0.102 | r=0.302<br>p=0.295 | **r=0.331**<br>**p=0.002** | **r=0.366**<br>**p=0.001** | **r=0.222**<br>**p=0.045** | **r=0.227**<br>**p=0.040** |

Neither IgG anti-HDL nor IgG anti-ApoA1 were associated with disease activity in RA patients (DAS28: r=-0.096, p=0.395 and r=0.091, p=0.418; SDAI: r=-0.109, p=0.332 and r=0.132, p=0.239, respectively). No correlations were found in other clinical features such as symptoms duration, morning stiffness or acute-phase reactant levels (all p<0.050). Equivalent findings were observed in CSA individuals, although IgG anti-ApoA1 were positively associated with ESR (r=0.670, p=0.013) in this group. The levels of IgG anti-HDL or anti-ApoA1 were not influenced by RF (RA: p=0.661 and p=0.836, CSA: p=0.491 and p=0.999, respectively) or ACPA positivity (RA: p=0.616 and p=0.852, CSA: p=0.259 and p=0.620, respectively). Furthermore, traditional CV risk factors were not associated with antibody levels (Supplementary Table 5).

Additionally, the associations between antibodies against HDL components and serum cytokines were examined. IgG anti-HDL levels were positively associated with IFNa, MIP- 1a, IL-6, IL-8 and IFNg, and a similar picture was found for their IgM counterparts, whereas a distinct pattern of associations was registered for anti-ApoA1 responses (Supplementary Table 6). In the CSA group, only IgM ApoA1 levels correlated with those of IL-12 (Supplementary Table 6).

Taken together, these findings revealed that different IgG, but not IgM, antibodies against HDL particles and ApoA1 were associated with unfavourable lipoprotein features in RA and CSA, respectively. A similar picture was observed with inflammatory mediators. Importantly, the levels of these antibodies were independent of disease features and traditional CV risk factors.

### IgG anti-HDL antibodies were associated with atherosclerosis burden and improved risk stratification in RA

Next, the associations between antibodies against HDL and subclinical atherosclerosis, alone or in combination with traditional CV risk factors, were analysed.

IgG anti-HDL levels were associated with plaque presence and number in RA patients, and equivalent findings were retrieved for IgG anti-ApoA1 in CSA (Table 2). When patients were stratified by mSCORE risk strata, IgG anti-HDL and anti-ApoA1 antibodies were related to atherosclerosis in the low-risk group (mSCORE<5) in RA (n=62, p=0.034) and CSA (n=13, p=0.019), respectively. No associations were observed for the IgM counterparts. Moreover, none of the antibodies was found to correlate cIMT or vascular stiffness in these groups (Table 2).

**Table 2:** Associations between antibodies against HDL and subclinical CVD features. Associations between levels of antibodies against HDL or ApoA1 and subclinical CVD features were analyzed by Spearman ranks’ tests or Mann-Withney U tests in CSA and RA groups. Coefficients (r) and p-values, or p-values for the difference between groups are shown. Those reaching statistical significance are highlighted in bold.

|  | CSA |  |  |  | RA |  |  |  |
| --- | --- | --- | --- | --- | --- | --- | --- | --- |
|  | Anti-HDL<br>IgG | Anti-HDL<br>IgM | Anti-ApoA1<br>IgG | Anti-ApoA1<br>IgM | Anti-HDL<br>IgG | Anti-HDL<br>IgM | Anti-ApoA1<br>IgG | Anti-ApoA1<br>IgM |
| *Subclinical atherosclerosis* |  |  |  |  |  |  |  |  |
| Plaque presence | p=0.148 | p=0.199 | **p=0.020** | p=0.503 | **p=0.012** | p=0.736 | p=0.116 | p=0.640 |
| Plaque number | r=0.461<br>p=0.113 | r=-0.420<br>p=0.154 | **r=0.650**<br>**p=0.016** | r=0.271<br>p=0.371 | **r=0.274**<br>**p=0.016** | r=0.057<br>p=0.622 | r=0.144<br>p=0.213 | r=0.074<br>p=0.522 |
| Plaque risk | p=0.215 | p=0.339 | p=0.319 | p=0.535 | p=0.535 | p=0.319 | p=0.339 | p=0.215 |
| cIMT | r=0.096<br>p=0.754 | r=0.465<br>p=0.109 | r=0.143<br>p=0.641 | r=0.033<br>p=0.915 | r=-0.031<br>p=0.791 | r=-0.214<br>p=0.061 | r=-0.025<br>p=0.830 | r=0.023<br>p=0.840 |
| *Vascular stiffness* |  |  |  |  |  |  |  |  |
| VS | r=0.414<br>p=0.205 | r=0.588<br>p=0.067 | r=0.030<br>p=0.931 | r=0.358<br>p=0.279 | r=0.155<br>p=0.282 | r=-0.052<br>p=0.722 | r=-0.146<br>p=0.312 | r=0.012<br>p=0.934 |
| VD | r=0.052<br>p=0.887 | r=0.309<br>p=0.386 | r=-0.057<br>p=0.875 | r=0.117<br>p=0.749 | r=0.064<br>p=0.676 | r=0.114<br>p=0.455 | r=-0.035<br>p=0.821 | r=0.019<br>p=0.902 |
| VSf | r=-0.013<br>p=0.971 | r=-0.276<br>p=0.441 | r=0.137<br>p=0.706 | r=-0.015<br>p=0.968 | r=-0.117<br>p=0.445 | r=-0.200<br>p=0.187 | r=-0.063<br>p=0.679 | r=-0.071<br>p=0.641 |
| PSEM | r=-0.014<br>p=0.912 | r=-0.376<br>p=0.322 | r=-0.439<br>p=0.237 | r=-0.324<br>p=0.395 | r=-0.147<br>p=0.334 | r=-0.220<br>p=0.147 | r=-0.055<br>p=0.721 | r=-0.091<br>p=0.552 |

In the RA group, those associations remained after adjusting for traditional CV risk factors as potential confounders (Table 3) (Supplementary Table 7). IgG anti-HDL levels alone were able to discriminate between patients with and without atherosclerosis (AUC [95% CI]: 0.669 [0.547–0.790], p=0.012). Adding IgG anti-HDL tertiles to the mSCORE (mSCORE + anti- HDL) improved the identification of RA patients with atherosclerosis (Table 4). Although adding those of IgG anti-ApoA1 led to certain improvement, superiority was demonstrated for anti-HDL resulting in a better discrimination capacity (difference between areas = 0.086 [0.023–0.150], p=0.007), improved classification metrics (sensibility, percentage of patients correctly classified, and Matthews Correlation coefficient) and risk prediction (Hosmer-Lemeshow statistic) (Table 4). NRI features clearly confirmed a better patient reclassification to higher risk categories for those presenting atherosclerosis with a negligible effect in those without. Furthermore, although achieving similar highest Youden indices, the optimal cut- off value achieved by adding IgG anti-HDL to the mSCORE was more realistic for stratification than that of mSCORE alone or adding anti-ApoA1 (Table 4), which was mostly specificity-skewed. Finally, IgG anti-ApoA1 levels were able to discriminate atherosclerosis status in CSA individuals (AUC: 0.819 [0.719–1.000], p=0.021), but the low sample size prevented multivariate analyses.

**Table 3:**
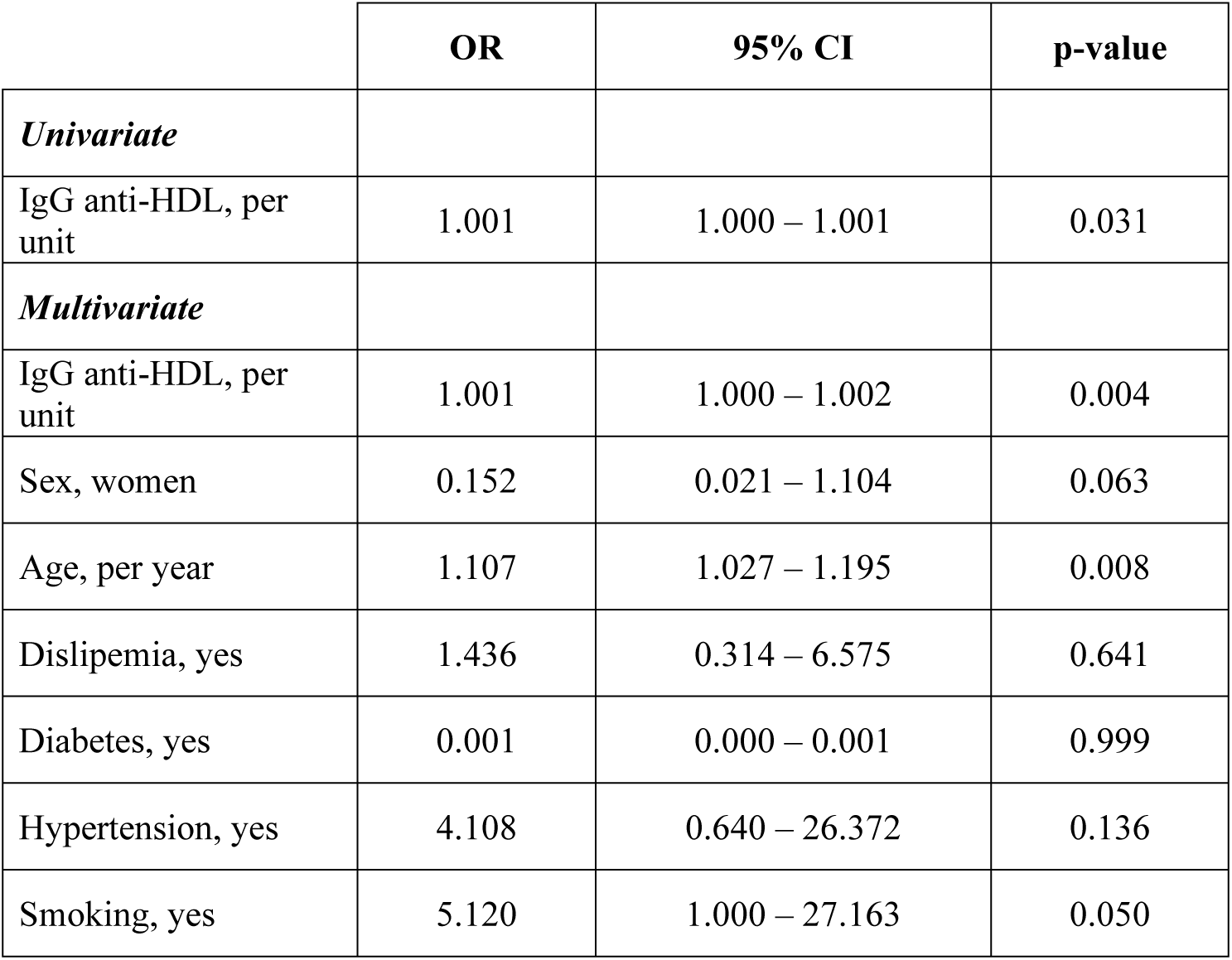
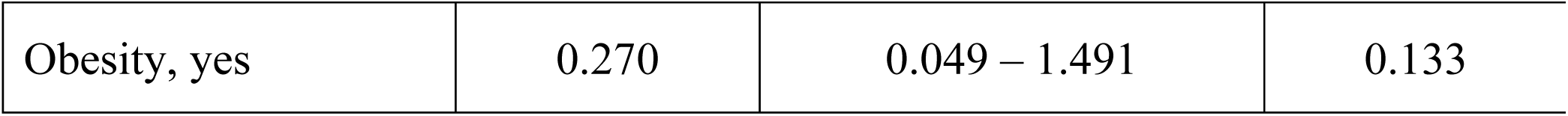
IgG anti-HDL as predictor of atherosclerosis plaque occurrence in RA. The role of IgG anti-HDL levels as predictor of atherosclerosis occurrence in early RA patients was analysed by univariate and multivariate logistic regression analyses. The presence of atherosclerosis plaque was entered as the dependent variable.

**Table 4:** IgG anti-HDL improved CV risk stratification in early RA. Analysis of the added value of IgG anti-HDL levels to the mSCORE risk stratification compared to the use of mSCORE alone or adding IgG anti-ApoA1 levels. Classification, calibration metrics and goodness-of-fit statistics are shown.

|  | **mSCORE** | **mSCORE + IgG anti-HDL** | **mSCORE + IgG anti-ApoA1** |
| --- | --- | --- | --- |
| AUC ROC (95% CI) | 0.636 (0.514 – 0.759) | 0.826 (0.731 – 0.922) | 0.740 (0.627 – 0.852) |
| p-value | p=0.044 | p<0.0001 | p=0.0003 |
| Mathews' Correlation Coefficient | 0.319<br>p=0.003 | 0.514<br>p<0.0001 | 0.431<br>p<0.001 |
| Hosmer-Lemeshow test | p=0.002 | p=0.207 | p<0.001 |
| R2 | 0.173 | 0.510 | 0.297 |
| OR (95% CI) | 13.12 (1.62 – 106.00) | 36.80 (7.67 – 176.93) | 10.500 (3.14 – 35.05) |
| % Patients Correctly Classified | 57.14 (45.37 – 68.19) | 80.52 (69.60 – 88.34) | 71.43 (59.83 – 80.86) |
| Likelihood Ratio (95% CI) | 9.43 (1.31 – 68.13) | 11.12 (2.87 – 43.01) | 4.72 (1.84 – 12.12) |
| Sensitivity | 30.43 (18.20 – 45.92) | 71.74 (56.32 – 83.54) | 60.87 (45.39 – 74.54) |
| Specificity | 96.77 (81.49 – 99.83) | 93.55 (77.16 – 98.87) | 87.10 (69.52 – 95.92) |
| Positive Predictive Value | 93.33 (66.03 – 99.65) | 94.29 (79.48 – 99.00) | 87.50 (70.07 – 95.92) |
| Negative Predictive Value | 48.39 (35.66 – 61.32) | 69.05 (52.76 – 81.69) | 60.00 (44.37 – 73.94) |
| Youden Index (value) | 0.632 (2.25) | 0.685 (4.75) | 0.634 (2.75) |
| NRI |  | 0.381 | 0.207 |
| NRI non-events |  | -0.032 | -0.096 |
| NRI events |  | 0.413 | 0.304 |

All these results that antibodies against HDL particles were independently associated with atherosclerosis burden in the earliest phases of arthritis. IgG anti-HDL levels improve patient stratification over conventional algorithms alone and were superior to their anti-ApoA1 counterparts.

### IgG anti-HDL response was associated with serum proteomic signatures related to immune activation, remodelling, and lipid metabolism

In order to get insight into the pathogenic mechanisms underlying the humoral responses against HDL components, the associations between antibody levels and serum proteomic profiles were evaluated in RA patients.

Several univariate correlations between proteomic features and IgG/IgM anti-HDL levels were detected (Supplementary Table 8). Some associations were also observed for IgG/IgM anti-ApoA1, although to a lower extent. After FDR controlling by Benjamini-Hochberg, a total of 23 features were associated with IgG anti-HDL, whereas 5 did with their IgM counterparts (Supplementary Table 9), and no associations were observed for anti-ApoA1 responses.

Proteins independently associated with IgG anti-HDL levels showed a significant protein-protein interaction enrichment (p<1.0·10^−16^) (Figure 2A) using the STRING platform. Protein nodes grouped into two main clusters, one including mostly immune and inflammatory mediators, and a second one including adhesion and extracellular matrix proteins, with PGF, ANGPT1, FGF21 and LPL located as hubs between clusters. Of note, some of these nodes showed major differences at the network level between patients with and without atherosclerosis (Supplementary Figure 3). Pathway annotation using ShinyGO uncovered functional pathways participated by these proteins, including immune activation, extracellular matrix homeostasis and remodelling, and response to cytokines (Figure 2B). Pathway analysis using KEGG mapper also identified other relevant pathways such as “cytokine-cytokine receptor interaction”, “rheumatoid arthritis”, “lipid and atherosclerosis” and “viral protein interaction with cytokine and cytokine receptor”. Finally, analyses by the TRRUST database identified nine candidate transcription factors that were shared for the proteins analyzed, thus underlining common expression programs (Supplementary Table 10).

**Figure 2:**
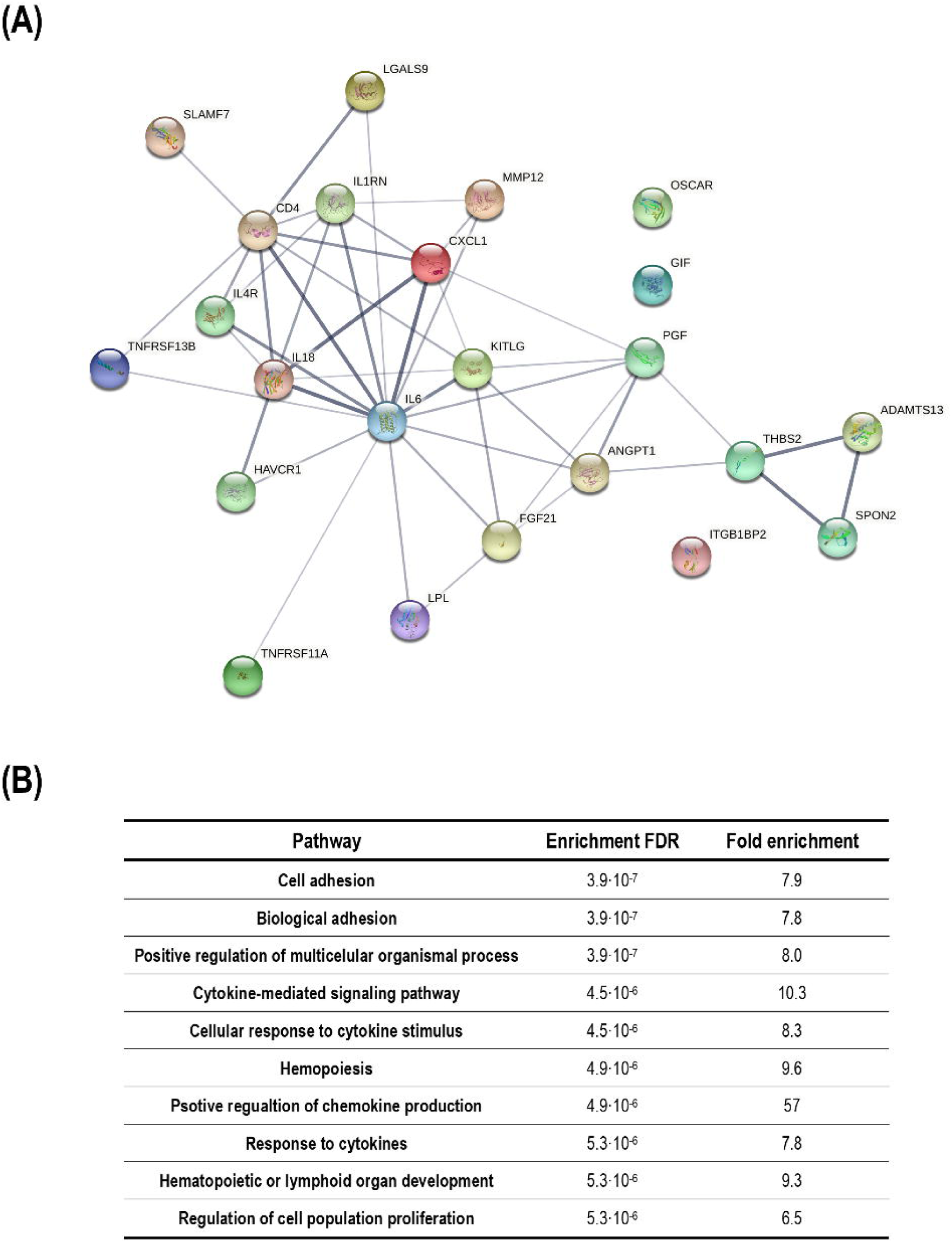
Pathogenic protein signatures related to IgG anti-HDL levels in early RA. (A) Protein-protein interactions among proteomic species found to be associated with IgG anti- HDL levels in early RA depicted in a network graph by the STRING platform. Two main clusters were identified. (B) Functional classification of the proteomic species into biological pathways (top 10) retrieved by the ShinyGO platform. Enrichment FDR and fold enrichment is indicated for each pathway identified.

These data suggest that different humoral responses against HDL exhibit distinct underlying serum proteome signatures, and IgG anti-HDL antibodies correlate with several proteins involved in pathogenic mechanisms related to immune activation, remodelling, and lipid metabolism in RA.

## DISCUSSION

The role of the humoral response as the missing link between autoimmunity, lipoproteins and CVD has gained attention in recent years, especially in the field of systemic autoimmune rheumatic diseases. Herein we demonstrated that humoral responses against HDL particles are an early event within RA disease course, although quantitative and qualitative differences can be noticed among stages. These differences were paralleled by distinct capacities for improving risk stratification, as well as with associations with lipoprotein particle size, content, functionality, and with underlying pathogenic pathways.

A major breakthrough of this study is the characterization of the antibody responses against HDL particles during the earliest phases of inflammatory arthritis. Our findings confirmed that antibodies against HDL and its components were not only present already at disease onset, but also before the clinical diagnosis can be established. Interestingly, during the arthralgia stage only the IgG response against ApoA1 was significantly increased and a strong correlation with that of against HDL was noted, hence suggesting that all anti-HDL response is mostly anti-ApoA1-directed. On the contrary, this association was much weaker in the clinical phase of the disease, thus pointing to the emergence of other specificities within the anti-HDL response around disease diagnosis. Of note, the responses were comparable between the early and established stages, thus suggesting that the repertoire is stable after disease onset, even despite exposure to disease duration and treatments. Therefore, these findings mirror those reported for the ACPA/RF trends along disease course in RA (20,21). Of note, the differences in specificities herein reported were also associated with clinical (CVD-related) outcomes, hence expanding the relevance of the ‘epitope spreading’ phenomenon (21) not only immunologically (beyond ACPA/RF), but also clinically (beyond arthritis onset). Taken together, these results strengthen the notion that CV-related alterations appear very early in the RA course in a subset of patients and follow a parallel progression, presumably by sharing pathogenic mechanisms, with other disease manifestations. Due to their early emergence around disease onset, whether they have prognostic properties warrants further studies.

A remarkable result was the comparative analysis of IgG anti-HDL and anti-ApoA1 responses. Until date, few comparative studies have been published, and the literature seems to be shifted towards ApoA1-targeted approaches, although supportive empirical evidence is scarce. Our findings shed new light into this topic. Contrary to what may be expected, both antibodies were only mildly correlated, especially in clinical disease. This is in line with reports by other authors in other conditions (16). This poor correlation led to important differences in clinical significance, where IgG anti-HDL demonstrated to be superior in RA. Two, non-exclusive, main hypotheses may explain this finding. First, it must be noted that HDL are complex structures with a substantial and diverse protein cargo, including several inflammatory mediators (22). The vasculo-protective functions are thus carried out by a range of different proteins. Anti-HDL responses may block different molecules, hence simultaneously counteracting several HDL activities and causing a strong, multi-level HDL dysfunction, which is more likely to cause an effect at the clinical level. This aligns with the associations observed with lipoprotein particle size distribution and content, as well as with the PON1 activity. Of note, these features are known to play a much more important role in atheroprotection than circulating HDL-C levels. Second, RA and other rheumatic conditions are hallmarked by the lipid paradox (3). Inflammation is known to both reduce HDL-C levels, but also to trigger changes on its protein composition (23,24), mostly by increasing acute-phase reactants and decreasing ApoA1 abundance (25–28). In fact, anti-ApoA1 antibodies have been reported to fluctuate in lupus patients (29), and the correlation between anti-HDL and anti-PON1 seems to depend on disease activity in RA (30). Similarly, anti- PON1 antibodies have demonstrated to account for a larger proportion of anti-HDL variance than anti-ApoA1 in psoriasis (31), despite the difference in abundance of these protein targets. However, the significance of anti-PON1 antibodies in RA is limited compared to that of anti-HDL (30). Therefore, it is tempting to speculate that reducing the analyses of the humoral response against lipoproteins to a single antigen, even more if it is ApoA1, may be too simplistic especially under high-grade inflammatory conditions. This may account for the lack of associations between anti-ApoA1 responses and CV outcomes in a number of conditions (32,33), including lupus patients (29,34). In fact, only a modest effect has been observed in established RA patients (35). Understanding the diversity of antibodies binding HDL particles may bring new clues for patient stratification and potential novel pathogenic mediators. Consequently, our data reinforce the need of considering anti-HDL responses as the standard in this scenario. However, and also balancing technical and experimental requirements, the use of anti-ApoA1 responses may be considered for certain, specific conditions, where inflammation is mildly or low-grade involved. In fact, results with anti- ApoA1 in CSA were comparable to those on anti-HDL in RA, hence strengthening this notion. This may also account for the added value of these autoantibodies in other scenarios (36–38), although a comparative analyses with that of anti-HDL are almost lacking in the literature.

Given the differences in added clinical value between these autoantibodies, we then investigated the underlying pathogenetic circuits to get insight into potential mechanistic pathways. First, protein signatures differed between IgG and IgM responses against the same target, thus stressing the relevance of class-switching and response maturation for their potential functional correlates. Our serum proteomic study coupled with a functional enrichment analysis confirmed that IgG anti-HDL, but not anti-ApoA1, response was associated with an enhanced pro-inflammatory milieu, elevated vascular and extracellular matrix turnover, cell adhesion and lipid metabolism. Importantly, all these biological processes are central to atherosclerosis occurrence and progression (39). Furthermore, no associations were found with anti-ApoA1 responses, hence underlining the relevance of other antigenic targets within the HDL structure in relation to their functional correlates. The involvement of some of the inflammatory mediators (such as IFNa, IFNg, IL-6, IL-8, TNF superfamily-related, etc) have been described in established disease by our group (12) and others (40), thus confirming these connections and strengthen their relevance in the early stage. Other proteins are indicative of shared mechanisms between joint and vascular involvement (such as hOSCAR, TNF superfamily members, ADAMTS13, etc); as well as interactions between inflammatory pathways and adipocyte tissue and glucose metabolism (FGF21). The association between anti-HDL and LPL levels is remarkable, as the latter is of major relevance as a key regulator of the inflammation/lipid metabolism axis. However, its involvement in RA is far from being clear (41). The positive correlation between anti-HDL and LPL may explain the association between the former and the lipoprotein triglyceride content observed in our study, since reduced LPL has been linked to reduced lipolysis and triglyceride clearance (42). Of note, diminished LPL levels have been described to associate with unfavourable lipid profiles and represent a risk factor itself (43,44). Therefore, the association between anti-HDL and LPL may account for the triglyceride-rich lipoproteins and cholesterol remnant accumulation in RA, which has been already reported elsewhere but underlying causes are unclear (45–47). Moreover, our proteomic approach revealed the existence of strong protein-protein interactions, which are related to anti-HDL responses and differ between patients with and without atherosclerosis. This is also supported by the observation of common transcription factors identified in our analyses. In view of these shared expression programs, it may be conceivable to analyze whether these protein hubs represent novel therapeutic targets that may be actionable by existing or experimental drugs.

Interestingly, the levels of anti-HDL or anti-ApoA1 were unrelated to traditional CV risk factors. On the one hand, this poses into question the use of algorithms solely based on these risk factors, which may explain why conventional algorithms underperform risk stratification. On the other hand, this may be responsible for the clinical added value observed in our analysis, especially for anti-HDL antibodies. The addition of these antibodies to the mSCORE resulted in a significant change in the goodness of fit, sensitivity and frequency of patients correctly classified into appropriate risk groups between the reference and the new models including the antibodies. The same applies between the anti-HDL-containing model and that of anti-ApoA1, again reinforcing the role of other antigenic targets. A similar conclusion has been reached by other authors, even in non-autoimmune disorders (38). Although there are some studies confirming that anti-ApoA1 improves risk stratification in some conditions over conventional algorithms (40), unfortunately comparative analyses with anti-HDL are very limited. Importantly, autoantibodies against lipoproteins have demonstrated their robustness as biomarkers compared to other soluble species (48). Therefore, our findings demonstrate the clinical potential of these mediators and their ability to cover important clinical unmet needs included in the research agenda for cardiovascular management proposed by EULAR (49). Additionally, due to the absence of validated clinical assays for HDL functionality, measurement of IgG anti-HDL levels may provide an indirect estimation in this setting. Since anti-HDL emergence is a common hallmark in a wide range of rheumatic conditions, it is tempting to speculate that these results may be of interest beyond RA, where similar research needs have been detected (50).

In conclusion, antibodies against HDL components are present in the earliest phases of RA, and relate to lipoprotein particle size and content, antioxidant functionality, inflammatory milieu and subclinical atherosclerosis burden, but not with traditional CV risk factors. IgG anti-HDL antibodies improve risk stratification in RA patients and correlate with several pathogenic pathways involved in atherosclerosis development. To the best of our knowledge, this is the first study characterizing the humoral response against HDL in the early stages of arthritis as well as in demonstrating the anti-HDL added clinical value. Our study has some limitations such as cross-sectional design and lack of follow-up although the association between anti-HDL and hard clinical endpoints has already been demonstrated by our group. Prospective studies are required to assess potential differences in prognostic value of anti- HDL and anti-ApoA1.

## Data Availability

All data produced in the present work are contained in the manuscript

## Author contributions

All authors were involved in drafting the manuscript or revising it critically for important intellectual content and all the authors gave their approval of the final version of the manuscript to be published.

Study conception and design: JRC, AS

Acquisition of data: JRC, MAL, PL, AIPA, SAC, NA, AS

Analysis and interpretation of data: JRC, MAL, GAR, FA, AS

## Funding

This work was supported by European Union FEDER funds, Fondo de Investigación Sanitaria from “Instituto de Salud Carlos III (ISCIII)” (reference PI21/00054), the Intramural Program from “Instituto de Investigación Sanitaria del Principado de Asturias (ISPA)” (reference 2021-046-INTRAMURAL NOV-ROCAJ), and a grant (reference Q122RSV03) from the European Alliance of Associations for Rheumatology (EULAR). The content is solely the responsibility of the authors and does not necessarily represent the official views of EULAR

## Competing interests

The authors declare that the research was conducted in the absence of any commercial or financial relationships that could be construed as a potential conflict of interest. Dr. Amigó has a patent method for lipoprotein characterization licensed to Biosfer Teslab (Spain) from which is stock owner, a company that commercialize the lipoprotein profiles described in the present manuscript. The funders had no role in study design, data analysis, interpretation, or decision to publish.

## Ethics approval

The study was approved by the local institutional review board (Comité de Ética de Investigación Clínica del Principado de Asturias) in compliance with the Declaration of Helsinki (reference CEImPA 2021.126). All study subjects gave written informed consent.

---

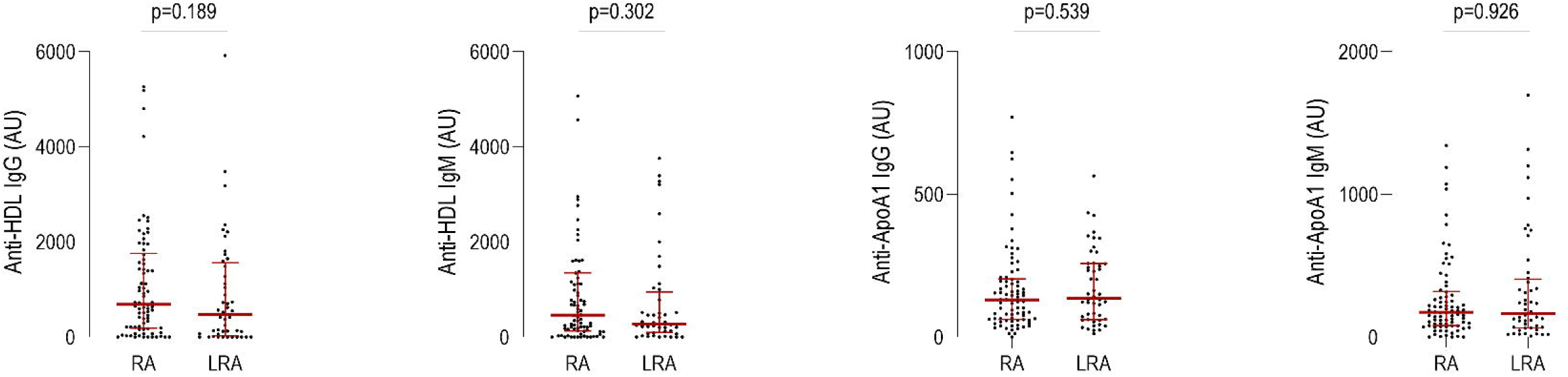

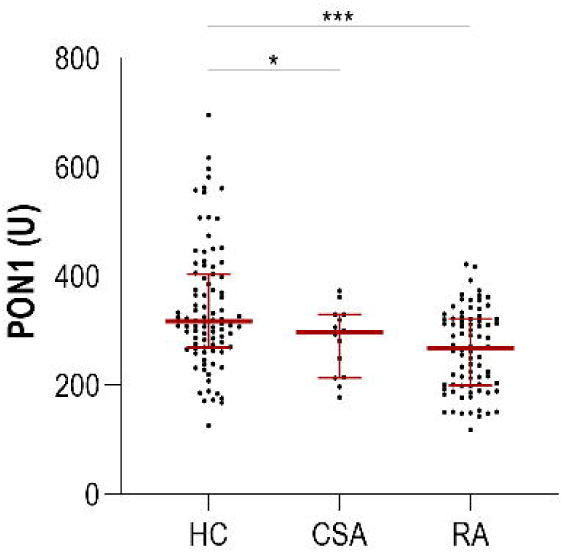

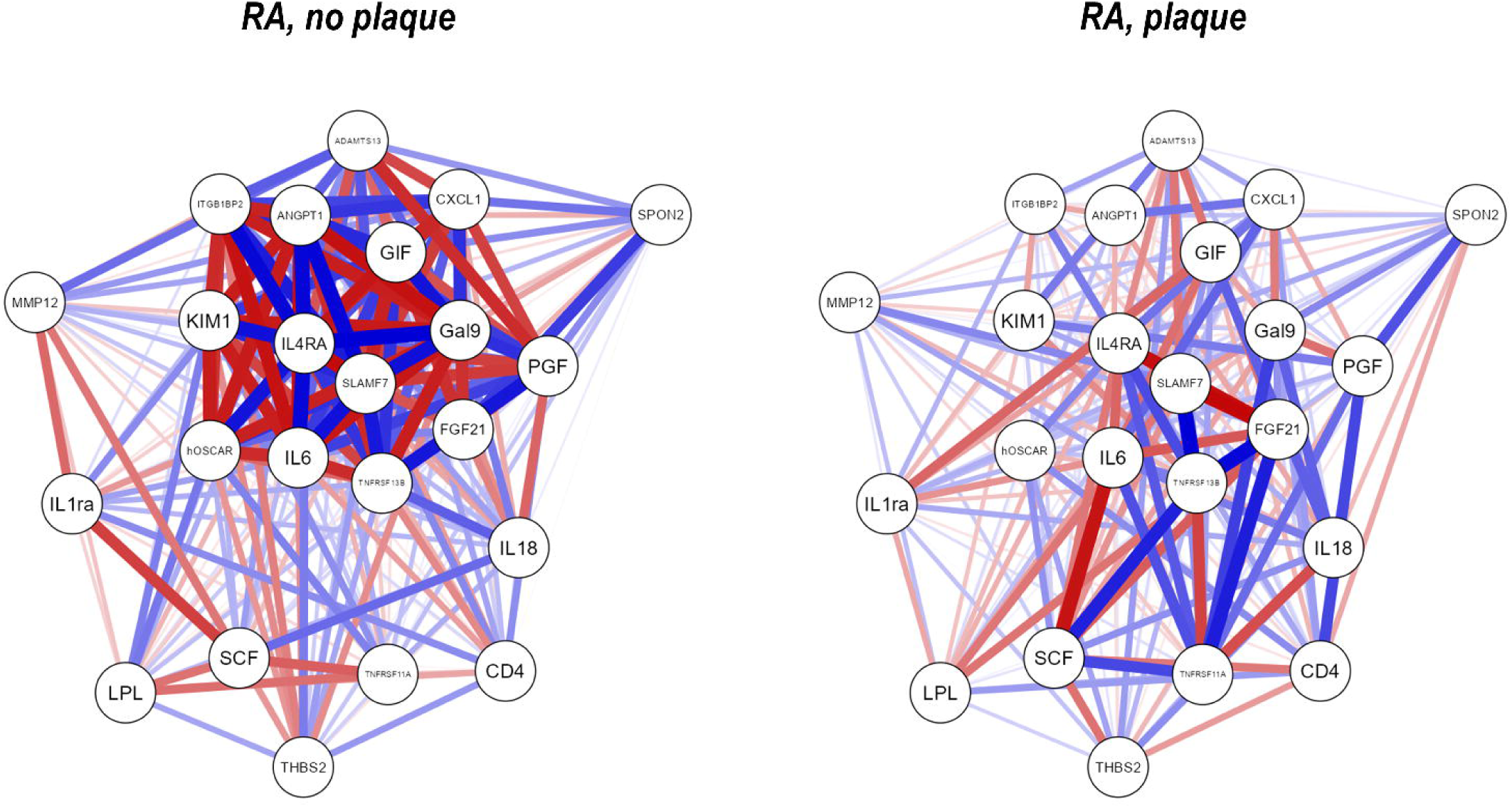

## Notes

### Competing Interest Statement

The authors have declared no competing interest.

### Funding Statement

This work was supported by European Union FEDER funds, Fondo de Investigacion Sanitaria from Instituto de Salud Carlos III (ISCIII) (reference PI21/00054), the Intramural Program from Instituto de Investigacion Sanitaria del Principado de Asturias (ISPA) (reference 2021-046-INTRAMURAL NOV-ROCAJ), and a grant (reference Q122RSV03) from the European Alliance of Associations for Rheumatology (EULAR). The content is solely the responsibility of the authors and does not necessarily represent the official views of EULAR.

### Author Declarations

The study was approved by the local institutional review board (Comite de Etica de Investigacion Clinica del Principado de Asturias) in compliance with the Declaration of Helsinki (reference CEImPA 2021.126). All study subjects gave written informed consent.

## REFERENCES

1. Symmons DPM, Gabriel SE. Epidemiology of CVD in rheumatic disease, with a focus on RA and SLE. Nature Reviews Rheumatology 2011;7:399–408. Available at: http://www.nature.com/articles/nrrheum.2011.75.

2. Rincón ID del, Williams K, Stern MP, Freeman GL, Escalante A. High incidence of cardiovascular events in a rheumatoid arthritis cohort not explained by traditional cardiac risk factors. Arthritis Rheum 2001;44:2737–45. Available at: http://www.ncbi.nlm.nih.gov/pubmed/11762933.

3. Myasoedova E, Crowson CS, Kremers HM, Roger VL, Fitz-Gibbon PD, Therneau TM, et al. Lipid paradox in rheumatoid arthritis: the impact of serum lipid measures and systemic inflammation on the risk of cardiovascular disease. Ann Rheum Dis 2011;70:482–7. Available at: http://www.ncbi.nlm.nih.gov/pubmed/21216812.

4. März W, Kleber ME, Scharnagl H, Speer T, Zewinger S, Ritsch A, et al. HDL cholesterol: reappraisal of its clinical relevance. Clinical Research in Cardiology 2017;106:663–675.

5. Robertson J, Peters MJ, McInnes IB, Sattar N. Changes in lipid levels with inflammation and therapy in RA: a maturing paradigm. Nat Rev Rheumatol 2013;9:513–23. Available at: http://www.ncbi.nlm.nih.gov/pubmed/23774906.

6. Navab M, Reddy ST, Lenten BJ van, Fogelman AM. HDL and cardiovascular disease: atherogenic and atheroprotective mechanisms. Nature Reviews Cardiology 2011;8:222–232.

7. Watanabe J, Charles-Schoeman C, Miao Y, Elashoff D, Lee YY, Katselis G, et al. Proteomic profiling following immunoaffinity capture of high-density lipoprotein: association of acute-phase proteins and complement factors with proinflammatory high-density lipoprotein in rheumatoid arthritis. Arthritis Rheum 2012;64:1828–37. Available at: http://www.ncbi.nlm.nih.gov/pubmed/22231638.

8. Choy E, Sattar N. Interpreting lipid levels in the context of high-grade inflammatory states with a focus on rheumatoid arthritis: a challenge to conventional cardiovascular risk actions. Ann Rheum Dis 2009;68:460–9. Available at: http://www.ncbi.nlm.nih.gov/pubmed/19286905.

9. Navab M, Reddy ST, Lenten BJ van, Anantharamaiah GM, Fogelman AM. The role of dysfunctional HDL in atherosclerosis. J Lipid Res 2009;50 Suppl:S145–S149.

10. Rodríguez-Carrio J, López-Mejías R, Alperi-López M, López P, Ballina-García FJ, González-Gay MÁ, et al. Paraoxonase 1 Activity Is Modulated by the rs662 Polymorphism and IgG Anti-High-Density Lipoprotein Antibodies in Patients With Rheumatoid Arthritis: Potential Implications for Cardiovascular Disease. Arthritis & Rheumatology 2016;68:1367–1376.

11. López P, Rodríguez-Carrio J, Martínez-Zapico A, Pérez-Álvarez Á, López-Mejías R, Benavente L, et al. Serum Levels of Anti-PON1 and Anti-HDL Antibodies as Potential Biomarkers of Premature Atherosclerosis in Systemic Lupus Erythematosus. Thrombosis and Haemostasis 2017;117:2194–2206. Available at: http://www.thieme-connect.de/DOI/DOI?10.1160/TH17-03-0221.

12. Rodríguez-Carrio J, Alperi-López M, López P, Ballina-García FJ, Abal F, Suárez A. Antibodies to high-density lipoproteins are associated with inflammation and cardiovascular disease in rheumatoid arthritis patients. Translational Research 2015;166:529–539. Available at: http://linkinghub.elsevier.com/retrieve/pii/S1931524415002522.

13. Rodríguez-Carrio J, Mozo L, López P, Nikiphorou E, Suárez A. Anti-High-Density Lipoprotein Antibodies and Antioxidant Dysfunction in Immune-Driven Diseases. Frontiers in Medicine 2018;5. Available at: http://journal.frontiersin.org/article/10.3389/fmed.2018.00114/full.

14. Delgado Alves J, Ames PRJ, Donohue S, Stanyer L, Nourooz-Zadeh J, Ravirajan C, et al. Antibodies to high-density lipoprotein and beta2-glycoprotein I are inversely correlated with paraoxonase activity in systemic lupus erythematosus and primary antiphospholipid syndrome. Arthritis Rheum 2002;46:2686–94. Available at: http://www.ncbi.nlm.nih.gov/pubmed/12384928.

15. Batuca JR, Ames PRJ, Amaral M, Favas C, Isenberg DA, Delgado Alves J. Anti-atherogenic and anti-inflammatory properties of high-density lipoprotein are affected by specific antibodies in systemic lupus erythematosus. Rheumatology (Oxford) 2009;48:26–31. Available at: http://www.ncbi.nlm.nih.gov/pubmed/19000993.

16. O’Neill SG, Giles I, Lambrianides A, Manson J, D’Cruz D, Schrieber L, et al. Antibodies to apolipoprotein A-I, high-density lipoprotein, and C-reactive protein are associated with disease activity in patients with systemic lupus erythematosus. Arthritis and Rheumatism 2010;62:845–854.

17. Ames P, Matsuura E, Batuca J, Ciampa A, Lopez L, Ferrara F, et al. High-density lipoprotein inversely relates to its specific autoantibody favoring oxidation in thrombotic primary antiphospholipid syndrome. Lupus 2010;19:711–716. Available at: http://journals.sagepub.com/doi/10.1177/0961203309357765.

18. Nielen MMJ. Simultaneous development of acute phase response and autoantibodies in preclinical rheumatoid arthritis. Annals of the Rheumatic Diseases 2006;65:535–537. Available at: http://ard.bmj.com/cgi/doi/10.1136/ard.2005.040659.

19. Nielen MMJ, Schaardenburg D van, Reesink HW, Stadt RJ van de, Horst-Bruinsma IE van der, Koning MHMT de, et al. Specific autoantibodies precede the symptoms of rheumatoid arthritis: A study of serial measurements in blood donors. Arthritis & Rheumatism 2004;50:380–386. Available at: http://doi.wiley.com/10.1002/art.20018.

20. Sokolove J, Bromberg R, Deane KD, Lahey LJ, Derber LA, Chandra PE, et al. Autoantibody Epitope Spreading in the Pre-Clinical Phase Predicts Progression to Rheumatoid Arthritis. PLoS ONE 2012;7:e35296.

21. Woude D van der, Rantapaa-Dahlqvist S, Ioan-Facsinay A, Onnekink C, Schwarte CM, Verpoort KN, et al. Epitope spreading of the anti-citrullinated protein antibody response occurs before disease onset and is associated with the disease course of early arthritis. Annals of the Rheumatic Diseases 2010;69:1554–1561.

22. Heinecke JW. The HDL proteome: a marker–and perhaps mediator–of coronary artery disease. Journal of Lipid Research 2009;50:S167–S171.

23. Shao B, Pennathur S, Pagani I, Oda MN, Witztum JL, Heinecke JW, et al. Modifying Apolipoprotein A-I by Malondialdehyde, but Not by an Array of Other Reactive Carbonyls, Blocks Cholesterol Efflux by the ABCA1 Pathway. Journal of Biological Chemistry 2010;285:18473–18484. Available at: https://linkinghub.elsevier.com/retrieve/pii/S0021925819575561.

24. Vaisar T, Pennathur S, Green PS, Gharib SA, Hoofnagle AN, Cheung MC, et al. Shotgun proteomics implicates protease inhibition and complement activation in the antiinflammatory properties of HDL. Journal of Clinical Investigation 2007;117:746–756. Available at: http://www.jci.org/cgi/doi/10.1172/JCI26206.

25. Rohrer L, Hersberger M, Eckardstein A von. High density lipoproteins in the intersection of diabetes mellitus, inflammation and cardiovascular disease. Current Opinion in Lipidology 2004;15:269–278.

26. Zheng L, Nukuna B, Brennan M-L, Sun M, Goormastic M, Settle M, et al. Apolipoprotein A-I is a selective target for myeloperoxidase-catalyzed oxidation and functional impairment in subjects with cardiovascular disease. Journal of Clinical Investigation 2004;114:529–541.

27. Hedrick CC, Thorpe SR, Fu M-X, Harper CM, Yoo J, Kim S-M, et al. Glycation impairs high-density lipoprotein function. Diabetologia 2000;43:312–320.

28. Fisher EA, Feig JE, Hewing B, Hazen SL, Smith JD. High-Density Lipoprotein Function, Dysfunction, and Reverse Cholesterol Transport. Arteriosclerosis, Thrombosis, and Vascular Biology 2012;32:2813–2820.

29. Nigolian H, Ribi C, Courvoisier DS, Pagano S, Alvarez M, Trendelenburg M, et al. Anti-apolipoprotein A-1 autoantibodies correlate with disease activity in systemic lupus erythematosus. Rheumatology (Oxford) 2020;59:534–544.

30. Rodríguez-Carrio J, Alperi-López M, López-Mejías R, López P, Ballina-García FJ, Abal F, et al. Antibodies to paraoxonase 1 are associated with oxidant status and endothelial activation in rheumatoid arthritis. Clinical Science 2016;130:1889–1899.

31. Paiva-Lopes MJ, Batuca JR, Gouveia S, Alves M, Papoila AL, Alves JD. Antibodies towards high-density lipoprotein components in patients with psoriasis. Arch Dermatol Res 2020;312:93–102.

32. Pruijm M, Schmidtko J, Aho A, Pagano S, Roux-Lombard P, Teta D, et al. High Prevalence of Anti-Apolipoprotein/A-1 Autoantibodies in Maintenance Hemodialysis and Association With Dialysis Vintage. Therapeutic Apheresis and Dialysis 2012;16:588–594. Available at: https://onlinelibrary.wiley.com/doi/10.1111/j.1744-9987.2012.01102.x.

33. Rubini Gimenez M, Pagano S, Virzi J, Montecucco F, Twerenbold R, Reichlin T, et al. Diagnostic and prognostic value of autoantibodies anti-apolipoprotein A-1 and anti-phosphorylcholine in acute non-ST elevation myocardial infarction. European Journal of Clinical Investigation 2015;45:369–379. Available at: https://onlinelibrary.wiley.com/doi/10.1111/eci.12411.

34. Croca S, Bassett P, Chambers S, Davari M, Alber K, Leach O, et al. IgG anti-apolipoprotein A-1 antibodies in patients with systemic lupus erythematosus are associated with disease activity and corticosteroid therapy: an observational study. Arthritis Research & Therapy 2015;17:26. Available at: http://arthritis-research.com/content/17/1/26.

35. Vuilleumier N, Bratt J, Alizadeh R, Jogestrand T, Hafström I, Frostegård J. Anti-apoA-1 IgG and oxidized LDL are raised in rheumatoid arthritis (RA): potential associations with cardiovascular disease and RA disease activity. Scandinavian Journal of Rheumatology 2010;39:447–453. Available at: http://www.tandfonline.com/doi/full/10.3109/03009741003742755.

36. Vuilleumier N, Montecucco F, Spinella G, Pagano S, Bertolotto M, Pane B, et al. Serum levels of anti-apolipoprotein A-1 auto-antibodies and myeloperoxidase as predictors of major adverse cardiovascular events after carotid endarterectomy. Thrombosis and Haemostasis 2013;109:706–715. Available at: http://www.thieme-connect.de/DOI/DOI?10.1160/TH12-10-0714.

37. Carbone F, Satta N, Montecucco F, Virzi J, Burger F, Roth A, et al. Anti-ApoA-1 IgG serum levels predict worse poststroke outcomes. European Journal of Clinical Investigation 2016;46:805–817. Available at: https://onlinelibrary.wiley.com/doi/10.1111/eci.12664.

38. Batuca J, Amaral M, Favas C, Justino G, Papoila A, Ames P, et al. Antibodies against HDL Components in Ischaemic Stroke and Coronary Artery Disease. Thrombosis and Haemostasis 2018;118:1088–1100. Available at: http://www.thieme-connect.de/DOI/DOI?10.1055/s-0038-1645857.

39. Libby P. The changing landscape of atherosclerosis. Nature 2021;592:524–533. Available at: http://www.nature.com/articles/s41586-021-03392-8.

40. Vuilleumier N, Bas S, Pagano S, Montecucco F, Guerne P-A, Finckh A, et al. Anti-apolipoprotein A-1 IgG predicts major cardiovascular events in patients with rheumatoid arthritis. Arthritis & Rheumatism 2010;62:2640–2650. Available at: https://onlinelibrary.wiley.com/doi/10.1002/art.27546.

41. Armas-Rillo L de, Quevedo-Abeledo JC, Hernández-Hernández V, Vera-González A de, González-Delgado A, García-Dopico JA, et al. The angiopoietin-like protein 4, apolipoprotein C3, and lipoprotein lipase axis is disrupted in patients with rheumatoid arthritis. Arthritis Research & Therapy 2022;24:99. Available at: https://arthritis-research.biomedcentral.com/articles/10.1186/s13075-022-02784-z.

42. Wang H, Eckel RH. Lipoprotein lipase: from gene to obesity. American Journal of Physiology-Endocrinology and Metabolism 2009;297:E271–E288. Available at: https://www.physiology.org/doi/10.1152/ajpendo.90920.2008.

43. Rip J, Nierman MC, Wareham NJ, Luben R, Bingham SA, Day NE, et al. Serum lipoprotein lipase concentration and risk for future coronary artery disease: the EPIC-Norfolk prospective population study. Arterioscler Thromb Vasc Biol 2006;26:637–42. Available at: http://www.ncbi.nlm.nih.gov/pubmed/16373616.

44. Kobayashi J, Nakajima K, Nohara A, Kawashiri M, Yagi K, Inazu A, et al. The relationship of serum lipoprotein lipase mass with fasting serum apolipoprotein B-48 and remnant-like particle triglycerides in type 2 diabetic patients. Hormone and metabolic research = Hormonund Stoffwechselforschung = Hormones et metabolisme 2007;39:612–6. Available at: http://www.ncbi.nlm.nih.gov/pubmed/17712727.

45. Burggraaf B, Breukelen-van der Stoep DF van, Zeben J van, Meulen N van der, Geijn G-JM van de, Liem A, et al. Evidence for increased chylomicron remnants in rheumatoid arthritis. European Journal of Clinical Investigation 2018;48:e12873. Available at: https://onlinelibrary.wiley.com/doi/10.1111/eci.12873.

46. Mena-Vázquez N, Rojas-Gimenez M, Jimenez Nuñez FG, Manrique-Arija S, Rioja J, Ruiz-Limón P, et al. Postprandial Apolipoprotein B48 is Associated with Subclinical Atherosclerosis in Patients with Rheumatoid Arthritis. Journal of Clinical Medicine 2020;9:2483. Available at: https://www.mdpi.com/2077-0383/9/8/2483.

47. Rodríguez-Carrio J, Alperi-López M, López P, López-Mejías R, Alonso-Castro S, Abal F, et al. High triglycerides and low high-density lipoprotein cholesterol lipid profile in rheumatoid arthritis: A potential link among inflammation, oxidative status, and dysfunctional high-density lipoprotein. Journal of Clinical Lipidology 2017;11:1043-1054.e2. Available at: https://linkinghub.elsevier.com/retrieve/pii/S1933287417303355.

48. Finckh A, Courvoisier DS, Pagano S, Bas S, Chevallier-Ruggeri P, Hochstrasser D, et al. Evaluation of cardiovascular risk in patients with rheumatoid arthritis: do cardiovascular biomarkers offer added predictive ability over established clinical risk scores? Arthritis Care Res (Hoboken) 2012;64:817–25. Available at: http://www.ncbi.nlm.nih.gov/pubmed/22302385.

49. Agca R, Heslinga SC, Rollefstad S, Heslinga M, McInnes IB, Peters MJL, et al. EULAR recommendations for cardiovascular disease risk management in patients with rheumatoid arthritis and other forms of inflammatory joint disorders: 2015/2016 update. Annals of the Rheumatic Diseases 2017;76:17–28. Available at: https://ard.bmj.com/lookup/doi/10.1136/annrheumdis-2016-209775.

50. Drosos GC, Vedder D, Houben E, Boekel L, Atzeni F, Badreh S, et al. EULAR recommendations for cardiovascular risk management in rheumatic and musculoskeletal diseases, including systemic lupus erythematosus and antiphospholipid syndrome. Annals of the Rheumatic Diseases 2022;81:768–779. Available at: https://ard.bmj.com/lookup/doi/10.1136/annrheumdis-2021-221733.

