## Supplementary Materials for "Humoral responses against HDL particles are linked to lipoprotein traits, atherosclerosis occurrence, inflammation and pathogenic pathways during the earliest stages of arthritis"

**SUPPLEMENTARY MATERIAL**


| |
|---|
| **Supplementary Material and methods** |
| **Supplementary Tables** |
| **Supplementary Figure legends** |

### SUPPLEMENTARY MATERIAL AND METHODS

Study participants

A total of 82 early RA patients fulfilling the 2010 ACR/EULAR classification criteria (1) participated in this study. RA patients were recruited at disease onset from the early arthritic clinic of the Department of Rheumatology at Hospital Universitario Central de Asturias (HUCA), and therefore were not exposed to disease-modifying antirheumatic drugs (DMARDs) at the time of sampling. A complete clinical examination, including Disease Activity Score 28-joints (DAS28), Simplified Disease Activity Index (SDAI) and Health Assessment Questionnaire (HAQ) calculations, was carried out during the clinical appointment.

Subjects with clinically-suspect arthralgia (CSA) (n=14) were recruited from the same clinic if they met at least four criteria of the EULAR definition of arthralgia suspicious for progression to RA (2). A group of 96 healthy controls (HC) (70 women/26 men, mean age 60.00 (range 27.00 – 83.50) years) were recruited among age- and sex-matched healthy individuals from the same population.

A group (n=42) of long-lasting, established RA patients (LRA), also fulfilling 2010 ACR/EULAR classification criteria (1) but with more than 6 months of disease duration (median 6.76 years), was recruited from the same department as a validation cohort.

A fasting blood sample was collected from all individuals by venepuncture. Serum samples were immediately transferred to the laboratory and processed within less than 2 hours. Serum samples were stored at -80ºC until experimental procedures. Conventional blood biochemical (including CRP and ESR measurements) and lipid analyses were performed in all individuals. The presence of traditional CV risk factors (dyslipemia, hypertension, obesity, diabetes and smoking history) based on national guidelines was obtained from the medical records. Body measurements such as weight, height and waist circumference were performed at recruitment. CV risk was assessed using the mSCORE (3) and patients were grouped in risk categories (0-5: low and moderate; and >5: high and very high) according to the ESC consensus (4).

The study was approved by the local institutional review board (Comité de Ética de Investigación Clínica del Principado de Asturias) in compliance with the Declaration of

Helsinki (reference CEImPA 2021.126). All study subjects gave written informed consent.

Vascular imaging and functional assessments

Doppler ultrasound assessment was performed in the sonography laboratory (Department of Neurology, HUCA) by an experienced user blinded to the status of the study participants. All measures were carried out online in B-mode using a Toshiba Aplio XG device (Toshiba American Medical Systems) with software version 5.1. The carotid intima-media thickness (cIMT) was bilaterally measured (left and right common carotid arteries), according to the "Mannheim Carotid Intima-Media Thickness Consensus (2004-2006)" (5), and the mean value was used. The reproducibility of the cIMT measurement was 89.0%. Atherosclerotic plaque was defined as a distinct area protruding into the vessel lumen at least 0.5 mm, with 50% greater thickness than the cIMT found in surrounding areas or the presence of a cIMT>1.5 mm (5). Plaque prevalence was considered as the frequency of patients exhibiting at least one plaque according to the previous consensus definition. Plaque vulnerability was assessed by the ultrasound appearance of the plaques, and these were classified as "low" or "high" risk (6).

Vascular functionality was quantified by evaluating the diameter changes of the common carotid during an entire cardiac cycle. To this end, stiffness parameters (vascular strain and stiffness, vascular distensibility and pressure-strain elastic modulus (PSEM)) were calculated (7,8).

Quantification of antibodies against HDL particles

Levels of antibodies against HDL or ApoA1 were measured in serum samples by in-house enzyme-linked immunosorbent assays (ELISA).

Anti-HDL levels (both IgG and IgM) were measured as previously described (9) with slight modifications. First, Polysorp (Nunc) plates were half-coated overnight at room temperature with 20 μg/ml human HDL-cholesterol (Sigma) in 70% ethanol (test half) or ethanol alone (control half). The plates were blocked with 1% bovine serum albumin (BSA) in phosphate buffer saline (PBS) (pH 7.4) for 1 hour at 37º C and then washed with PBS. Next, standard curves (pooled sera diluted 1:16 to 1:512), diluted samples (1:50) were prepared in 0.1% BSA PBS and incubated for 2 h at room temperature under shacking. Unbound antibodies were removed by repeating the washing step. Alkaline phosphatase-conjugated anti-human IgG (1:1000) or IgM (1:500) (Immunostep) in 1% BSA tris buffer saline (TBS) were added and incubated for 1h. Finally, the plate was washed twice with TBS and 1 mg/ml p-nitrophenylphosphate (Sigma) diluted in dietanolamine buffer (pH 9.8) was added to the wells. Absorbance at 405 nm was recorded and the signal from the control half of the plate was subtracted to that of the test half. Anti-HDL Arbitrary Units (AU) were calculated for each sample according to the standard curves. Our assay had a within-run imprecision (coefficient of variation) of 9.81%, and a between-run coefficient of variation of 11%.

The protocol for anti-ApoA1 antibodies was adapted from the previous one, according to Frias et al. (10). Briefly, Polysorp plates were half-coated with 10 μg/ml human Apolipoprotein A1 (Sigma) in 70% ethanol (test half) or 70% ethanol alone (control half) for 2h at 37º C. Plates were blocked as previously described and incubated with standards and samples (diluted 1:100) for 2h at room temperature under shacking. Unbound antibodies were removed by washing with TBS and alkaline phosphatase-conjugated anti-human IgG (1:1000) or IgM (1:1000) were added in 1% BSA tris buffer saline (TBS) and incubated for 1h. The washing step was repeated, and substrate was added as previously described. Intra- and inter-assay coefficients of variation for anti-ApoA1 were 7.50% and 9.80%, respectively.

Assessment of PON1 activity

PON1 activity was quantified in serum samples by means of an enzymatic assay according to Eckerson et al. (11) with slight modifications, as previously reported (9). In brief, 7.5 μl of serum samples were incubated with 300 μl of 1 mM paraoxon (Sigma Aldrich) freshly prepared in 50 mM glycine buffer (with 1 mM $CaCl_2$, pH 10.5) in 96-well Maxisorp plates (Nunc) for 20 minutes at 37º C. The production of p-nitrophenol was monitored at 405 nm using a spectrophotometer. A unit (U) of PON1 activity was expressed as p-nitrophenol μmoles formed per minute and per ml of serum. Negative and quality controls were included in each plate to correct for inter-assay variations. Inter- and intra-assay coefficients of variation were 9.91% and 6.12%, respectively.

Lipoprotein characterization

An advanced lipoprotein characterization by means of the H-NMR-based Liposcale test (12) was performed. This protocol allowed the assessment of lipid content (cholesterol and triglycerides) of VLDL, IDL, LDL and HDL as well as the particle number and size (diameter) of VLDL LDL and HDL and their subclasses (small, medium and large). To this end, serum samples were diluted with deuterated water and 50 mM pH 7.4 phosphate buffer solution before H-NMR analysis. Then, H-NMR spectra were recorded at 310 K on a Bruker Advance III 600 spectrometer operating at a proton frequency of 600.20 MHz. Particle concentration and the diffusion coefficients were obtained from the measured amplitudes and attenuation of their spectroscopically distinct lipid methyl group NMR signals, using the 2D diffusion-ordered 1H NMR spectroscopy (DSTE) pulse as previously described (12).

Measurement of serum cytokine levels

The serum levels of IFNα and MIP1α were quantified using a Cytometric Bead Array Flex Set (BD) in a BD FACS Canto II flow cytometer using FCAP Array v.1.0.1, following the manufacturer's instructions. The detection limits were 1.25 pg/ml and 0.6 pg/ml, respectively.

IL-6, TNF, IFNγ, IL-1β, IL-23, IL-12 and IL-8 serum levels were assessed using a pre-defined multiplex assay (Human Inflmammation Panel 1, LEGENDplex, BioLegend) using the same device, according to the protocol provided by the manufacturer. The detection limits were 1.5 pg/ml, 0.9 pg/ml, 1.3 pg/ml, 1.5 pg/ml, 1.8 pg/ml, 2.0 pg/ml and 2.0 pg/ml, respectively.

Proteomic analysis

A pre-defined panel of 92 proteins related to CV (CV Panel II) was measured in serum samples using the Proximity Extension Assay proprietary test from Olink (Olink Bioscience, Sweden) (13).

Statistical analyses

Continuous variables were expressed as median (interquartile range) or mean±standard deviation, whereas categorical variables were summarized as n(%).

Differences among groups were evaluated by one-way ANOVA, Mann-Withney U, Kruskal-Wallis or $\chi 2$ tests, as appropriate. Correlations were assessed by Spearman ranks' tests. Corrections for multiple comparison tests were performed by Dunn-Bonferroni or by Benjamini-Hochberg procedure (14), as required. The associations between antibody levels and atherosclerosis occurrence were analysed by multiple logistic regression. Due to the overdispersion of the number of plaques, negative binomial regression models were used when studied as an outcome variable. Adjusted OR and 95% confidence intervals (CI) were calculated. The discrimination ability for subclinical CV disease was assessed using the area under the receiver operating characteristic curve (AUC ROC). In order to add the antibody levels to the mSCORE algorithm, categories were defined from the distribution of the antibody levels observed in the HC population using tertiles as cut-offs and a score was given as follows: 0 to T1: +0, T1 to T2: +1, and T2 to T3: +2 points. These points were added to the mSCORE values (mSCORE + anti-HDL, or mSCORE + anti-HDL).

The performance of the classification of mSCORE and mSCORE + antibodies was analysed by classification measures (sensitivity, specificity, % patients correctly classified and likelihood ratios), Matthews' correlation coefficient, goodness of fit (Hosmer–Lemeshow test) and the Youden index to determine the optimal cut-offs (threshold points with maximum accuracy). Differences in the AUC ROC values were compared by the De Jong statistic.

Proteomic data were evaluated by functional and pathway enrichment analyses using ShinyGo 0.76 (15) and KEGG platforms (16) to identify biological pathways. Protein-protein interactions were assessed by the STRING database (17). To investigate whether common transcriptional regulatory networks may underlie the protein networks, the TRRUST database (18) was used to predict transcription factors that regulate hub genes.

A p-value <0.050 was considered as statistically significant. Statistical analyses were carried out under SPSS v. 27, R v.4.1.3 and GraphPad Prism 8.0.

### REFERENCES


1. Aletaha D, Neogi T, Silman AJ, Funovits J, Felson DT, Bingham CO, et al. 2010 rheumatoid arthritis classification criteria: an American College of Rheumatology/European League Against Rheumatism collaborative initiative. *Ann Rheum Dis* 2010;69:1580–8. Available at: http://www.ncbi.nlm.nih.gov/pubmed/20699241.

2. Steenbergen HW van, Aletaha D, Beaart-van de Voorde LJJ, Brouwer E, Codreanu C, Combe B, et al. EULAR definition of arthralgia suspicious for progression to rheumatoid arthritis. *Annals of the Rheumatic Diseases* 2017;76:491–496. Available at: http://ard.bmj.com/lookup/doi/10.1136/annrheumdis-2016-209846.

3. Agca R, Heslinga SC, Rollefstad S, Heslinga M, McInnes IB, Peters MJL, et al. EULAR recommendations for cardiovascular disease risk management in patients with rheumatoid arthritis and other forms of inflammatory joint disorders: 2015/2016 update. *Ann Rheum Dis* 2017;76:17–28. Available at: http://www.ncbi.nlm.nih.gov/pubmed/27697765.

4. Piepoli MF, Hoes AW, Agewall S, Albus C, Brotons C, Catapano AL, et al. 2016 European Guidelines on cardiovascular disease prevention in clinical practice. *European Heart Journal* 2016;37:2315–2381.

5. Touboul P-J, Hennerici MG, Meairs S, Adams H, Amarenco P, Bornstein N, et al. Mannheim Carotid Intima-Media Thickness Consensus (2004–2006). *Cerebrovascular Diseases* 2007;23:75–80. Available at: https://www.karger.com/Article/FullText/97034.

6. Picano E, Paterni M. Ultrasound Tissue Characterization of Vulnerable Atherosclerotic Plaque. *International Journal of Molecular Sciences* 2015;16:10121–10133. Available at: http://www.mdpi.com/1422-0067/16/5/10121.

7. O'Rourke MF, Staessen JA, Vlachopoulos C, Duprez D, Plante G e. E. Clinical applications of arterial stiffness; definitions and reference values. *American Journal of Hypertension* 2002;15:426–444. Available at: https://academic.oup.com/ajh/article-lookup/doi/10.1016/S0895-7061(01)02319-6.

8. Zardi EM, Sambataro G, Basta F, Margiotta DPE, Afeltra AMV. Subclinical Carotid Atherosclerosis in Elderly Patients with Primary Sjögren Syndrome: A Duplex Doppler Sonographic Study. *International Journal of Immunopathology and Pharmacology* 2014;27:645–651. Available at: http://journals.sagepub.com/doi/10.1177/039463201402700422.

9. Rodríguez-Carrio J, Alperi-López M, López P, Ballina-García FJ, Abal F, Suárez A. Antibodies to high-density lipoproteins are associated with inflammation and cardiovascular disease in rheumatoid arthritis patients. *Translational Research* 2015;166:529–539. Available at: http://linkinghub.elsevier.com/retrieve/pii/S1931524415002522.

10. Frias MA, Virzi J, Batuca J, Pagano S, Satta N, Delgado Alves J, et al. ELISA methods comparison for the detection of auto-antibodies against apolipoprotein A1. *Journal of Immunological Methods* 2019;469:33–41. Available at: https://linkinghub.elsevier.com/retrieve/pii/S0022175919300407.

11. Eckerson HW, Wyte CM, Du BN la. The human serum paraoxonase/arylesterase polymorphism. *Am J Hum Genet* 1983;35:1126–38.

12. Mallol R, Amigó N, Rodríguez MA, Heras M, Vinaixa M, Plana N, et al. Liposcale: a novel advanced lipoprotein test based on 2D diffusion-ordered 1 H NMR spectroscopy. *Journal of Lipid Research* 2015;56:737–746. Available at: http://www.jlr.org/lookup/doi/10.1194/jlr.D050120.

13. Lundberg M, Eriksson A, Tran B, Assarsson E, Fredriksson S. Homogeneous antibody-based proximity extension assays provide sensitive and specific detection of low-abundant proteins in human blood. *Nucleic Acids Research* 2011;39:e102–e102. Available at: https://academic.oup.com/nar/article-lookup/doi/10.1093/nar/gkr424.

14. Benjamini Y, Hochberg Y. Controlling the False Discovery Rate: a Practical and Powerful Approach to Multiple testing. *J R Statist Soc B* 1995;57:289–300.

15. Ge SX, Jung D, Yao R. ShinyGO: a graphical gene-set enrichment tool for animals and plants. Valencia A, ed. *Bioinformatics* 2020;36:2628–2629. Available at: https://academic.oup.com/bioinformatics/article/36/8/2628/5688742.

16. Kanehisa M, Furumichi M, Sato Y, Ishiguro-Watanabe M, Tanabe M. KEGG: integrating viruses and cellular organisms. *Nucleic Acids Research* 2021;49:D545–D551. Available at: https://academic.oup.com/nar/article/49/D1/D545/5943834.

17. Szklarczyk D, Gable AL, Nastou KC, Lyon D, Kirsch R, Pyysalo S, et al. The STRING database in 2021: customizable protein–protein networks, and functional characterization of user-uploaded gene/measurement sets. *Nucleic Acids Research* 2021;49:D605–D612. Available at: https://academic.oup.com/nar/article/49/D1/D605/6006194.

18. Han H, Cho J-W, Lee S, Yun A, Kim H, Bae D, et al. TRRUST v2: an expanded reference database of human and mouse transcriptional regulatory interactions. *Nucleic Acids Research* 2018;46:D380–D386. Available at: http://academic.oup.com/nar/article/46/D1/D380/4566018.

### SUPPLEMENTARY TABLES

**Supplementary Table 1: Demographic, clinical and subclinical atherosclerosis assessments in RA and CSA.** Demographic, clinical features, traditional CV risk factors and subclinical atherosclerosis features of RA patients and CSA individuals were summarized. Variables were expressed as median (interquartile range), mean±SD or n(%), unless otherwise stated, according to the distribution of the variables.

| | **CSA n=14** | **RA n=82** |
|---|---|---|
| Age (years), mean (range) | 49.28 (30.00-62.33) | 58.51 (30.33-83.67) |
| Gender (female/male) | 14/0 | 66/16 |
| ***Clinical features*** | | |
| Duration of symptoms (weeks) | 24.00 (40.00) | 20.00 (22.00) |
| Morning stiffness (minutes) | 30.00 (50.00) | 60.00 (80.0) |
| Tender joint count | 3.00 (3.00) | 8.00 (7.00) |
| Swollen joint count | 0.00 (1.00) | 6.00 (5.00) |
| ESR (mm/h) | 7.50 (8.84) | 24.00 (27.00) |
| CRP (mg/dl) | 0.15 (0.30) | 0.80 (2.20) |
| Patient global assessment (VAS 0-100) | 30.00 (50.00) | 70.00 (25.00) |
| Pain assessment (VAS 0-10) | 5.00 (5.00) | 7.00 (2.00) |
| DAS28 | | 5.40 (1.78) |
| SDAI | | 24.22 (16.90) |
| HAQ | 0.55 (0.60) | 1.11 (1.00) |
| Fatigue (VAS 0-10) | 4.50 (5.00) | 5.00 (7.00) |
| RF+, n(%) | 8 (66.6) | 57 (69.5) |
| ACPA+, n(%) | 7 (58.3) | 56 (68.2) |
| ***Traditional CV risk factors*** | | |
| Hypertension, n(%) | 1 (12.5) | 28 (34.1) |
| Diabetes, n(%) | 0 (0.0) | 9 (10.9) |

| | | |
|---|---|---|
| Dyslipidemia, n(%) | 3 (37.5) | 24 (29.2) |
| Smoking, n(%) | 10 (71.4) | 31 (27.8) |
| Obesity (BMI>30 kg/$m^2$), n(%) | 3 (37.5) | 33 (40.2) |
| Waist circumference (cm) | 92.00 (16.00) | 101.00 (20.00) |
| ***Subclinical atherosclerosis (n=92)*** | n=13 | n=77 |
| Plaque presence, n(%) | 4 (30.7) | 46 (59.7) |
| Plaque number, mean (range) | 0.46 (0-3) | 0.96 (0-4) |
| High-risk plaque, n(%) | 0 (0.0) | 20 (25.9) |
| cIMT (mm) | 0.58±0.15 | 0.67±0.10 |
| VS | 0.10±0.03 | 0.10±0.04 |
| VD | 0.0055±0.0031 | 0.0044±0.002 |
| VSf | 504.24±286.37 | 540.04±239.61 |
| PSEM | 3.52±2.00 | 3.83±1.68 |

**Supplementary Table 2: Demographic and parameters of long-lasting, established RA (LRA).** Demographic and clinical features of established RA patients recruited as a validation cohort. Variables were expressed as median (interquartile range), mean±SD or n(%), unless otherwise stated, according to the distribution of the variables.

| | **LRA n=42** |
|---|---|
| Age (years), mean (range) | 52.58 (22.42-71.42) |
| Gender (female/male) | 36/6 |
| ***Clinical features*** | |
| Disease duration, mean (range) (years) | 6.76 (0.50 – 30.00) |
| Tender joint count | 2.00 (6.50) |
| Swollen joint count | 1.00 (4.00) |
| ESR (mm/h) | 13.00 (23.00) |
| CRP (mg/dl) | 0.50 (0.95) |
| Patient global assessment (VAS 0-100) | 45.00 (46.50) |
| Pain assessment (VAS 0-10) | 5.00 (4.50) |
| DAS28 | 3.46 (1.58) |
| HAQ | 1.00 (1.31) |
| RF+, n(%) | 19 (45.2) |
| ACPA+, n(%) | 23 (54.7) |
| ***Traditional CV risk factors*** | |
| Hypertension, n(%) | 15 (35.7) |
| Diabetes, n(%) | 2 (4.7) |
| Dyslipidemia, n(%) | 16 (38.0) |
| Smoking, n(%) | 15 (35.7) |
| Obesity (BMI>30 $kg/m^2$), n(%) | 10 (23.8) |
| ***Treatments, n(%)*** | |
| Glucocorticoids | 25 (59.5) |
| Methotrexate | 34 (80.9) |
| TNF blockers | 18 (42.8) |

| | |
|---|---|
| IL-6 blockers | 3 (19.0) |
| Leflunomide | 4 (9.5) |

**Supplementary Table 3: Lipoprotein characterization by H-NMR.** An advanced characterization of the lipoprotein profile was performed by H-NMR. Differences were assessed by a one-way ANOVA test. Variables were summarized as mean±SD.

| | HC | CSA | RA | p-value |
|---|---|---|---|---|
| ***Lipoprotein content*** | | | | |
| VLDL-C (mg/dl) | 10.39±7.06 | 13.07±7.64 | 17.50±12.78 | 0.116 |
| IDL-C (mg/dl) | 8.63±4.22 | 9.43±4.42 | 11.74±5.13 | 0.089 |
| LDL-C (mg/dl) | 134.78±25.16 | 133.51±26.67 | 136.62±30.79 | 0.739 |
| HDL-C (mg/dl) | 58.33±12.01 | 58.70±11.90 | 53.69±12.53 | 0.144 |
| VLDL-TG (mg/dl) | 56.95±27.18 | 60.90±32.89 | 78.94±63.97 | 0.222 |
| IDL-TG (mg/dl) | 9.91±3.92 | 10.04±3.49 | 12.07±3.77 | 0.055 |
| LDL-TG (mg/dl) | 13.12±3.64 | 15.67±4.34 | 17.79±5.44 | 0.079 |
| HDL-TG (mg/dl) | 15.57±4.01 | 15.28±5.99 | 16.63±4.81 | 0.277 |
| ***Lipoprotein subclases distribution*** | | | | |
| VLDL-P (nmol/l) | 40.99±21.02 | 44.20±24.48 | 57.90±43.04 | 0.208 |
| Large VLDL-P | 1.18±0.80 | 1.14±0.51 | 1.47±0.80 | 0.222 |
| Medium VLDL-P | 3.28±2.20 | 4.66±2.34 | 5.79±7.21 | 0.076 |
| Small VLDL-P | 37.06±20.19 | 38.36±21.84 | 50.63±36.11 | 0.207 |
| LDL-P (nmol/l) | 1377.57±251.19 | 1339.99±283.15 | 1400.82±321.86 | 0.655 |
| Large LDL-P | 188.88±37.76 | 188.17±33.89 | 189.20±37.76 | 0.871 |
| Medium LDL-P | 386.18±98.42 | 408.57±111.69 | 422.90±140.32 | 0.565 |
| Small LDL-P | 779.17±151.84 | 743.23±193.69 | 788.71±206.98 | 0.584 |
| HDL-P (μmol/l) | 29.08±4.60 | 29.51±4.36 | 27.78±5.25 | 0.179 |
| Large HDL-P | 0.25±0.03 | 0.26±0.03 | 0.27±0.04 | 0.510 |
| Medium HDL-P | 9.17±1.80 | 9.97±1.52 | 9.54±1.59 | 0.495 |
| Small HDL-P | 18.90±3.21 | 19.27±3.33 | 17.96±4.31 | 0.247 |
| ***Lipoprotein size*** | | | | |
| VLDL diameter (nm) | 41.95±0.56 | 42.18±0.34 | 42.03±0.42 | 0.133 |
| LDL diameter (nm) | 21.00±0.27 | 21.11±0.35 | 21.03±0.38 | 0.514 |
| HDL diameter (nm) | 8.11±0.06 | 8.27±0.06 | 8.29±0.10 | 0.512 |

**Supplementary Table 4: PON1 activity and lipoprotein profile.** Associations between serum PON1 activity and HDL features (particle number and size) were analysed by Spearman ranks' test in RA patients. Those reaching statistical significance are highlighted in bold.

| | **PON1** |
|---|---|
| ***Particle number*** | |
| HDL-P (mmol/l) | **r=0.275 p=0.016** |
| Large | r=0.046 p=0.682 |
| Medium | r=0.080 p=0.475 |
| Small | **r=0.314 p=0.004** |
| ***Particle diameter (mm)*** | |
| HDL-Z | **r=-0.276 p=0.012** |

**Supplementary Table 5: Associations between antibodies against HDL and traditional CV risk factors.** Associations between levels of antibodies against HDL or ApoA1 and traditional CV risk factors were analysed by Spearman's rank tests or Mann-Withney U tests in CSA and RA groups. Coefficients (r) and p-values, or p-values for the difference between groups are shown.

| | CSA | | | | RA | | | |
|---|---|---|---|---|---|---|---|---|
| | **Anti-HDL IgG** | **Anti-HDL IgM** | **Anti-ApoA1 IgG** | **Anti-ApoA1 IgM** | **Anti-HDL IgG** | **Anti-HDL IgM** | **Anti-ApoA1 IgG** | **Anti-ApoA1 IgM** |
| ***Demographic parameters*** | | | | | | | | |
| Age | r=-0.095<br>p=0.748 | r=-0.354<br>p=0.215 | r=-0.002<br>p=0.994 | r=0.374<br>p=0.185 | r=-0.028<br>p=0.804 | r=-0.088<br>p=0.434 | r=0.150<br>p=0.178 | r=0.006<br>p=0.954 |
| Sex | | | | | p=0.377 | p=0.925 | p=0.967 | p=0.208 |
| ***Traditional CV risk factors*** | | | | | | | | |
| Hypertension | p=0.999 | p=0.571 | p=0.714 | p=0.286 | p=0.728 | p=0.910 | p=0.273 | p=0.220 |
| Smoking | p=0.188 | p=0.733 | p=0.539 | p=0.839 | p=0.850 | p=0.059 | p=0.999 | p=0.059 |
| Dyslipemia | p=0.368 | p=0.368 | p=0.659 | p=0.659 | p=0.891 | p=0.659 | p=0.450 | p=0.453 |
| Diabetes | | | | | p=0.269 | p=0.953 | p=0.362 | p=0.045 |
| Obesity | p=0.263 | p=0.370 | p=0.172 | p=0.105 | p=0.393 | p=0.786 | p=0.786 | p=0.786 |

| | | | | | | | | |
|---|---|---|---|---|---|---|---|---|
| Waist circumference | r=0.321 p=0.482 | r=0.524 p=0.227 | r=0.332 p=0.467 | r=0.182 p=0.696 | r=0.009 p=0.941 | r=-0.002 p=0.985 | r=-0.114 p=0.350 | r=-0.180 p=0.139 |

**Supplementary Table 6: Associations between antibodies against HDL and serum cytokines.** Associations between levels of antibodies against HDL or ApoA1 and levels of serum cytokines were analysed by Spearman's rank tests in CSA and RA groups. Coefficients (r) and p-values are shown. Those reaching statistical significance are highlighted in bold.

| | CSA | | | | RA | | | |
|---|---|---|---|---|---|---|---|---|
| | **Anti-HDL IgG** | **Anti-HDL IgM** | **Anti-ApoA1 IgG** | **Anti-ApoA1 IgM** | **Anti-HDL IgG** | **Anti-HDL IgM** | **Anti-ApoA1 IgG** | **Anti-ApoA1 IgM** |
| IFNa | r=0.018<br>p=0.950 | r=0.120<br>p=0.683 | r=-0.167<br>p=0.568 | r=0.195<br>p=0.504 | **r=0.227<br>p=0.040** | **r=0.274<br>p=0.013** | r=0.072<br>p=0.523 | r=0.111<br>p=0.320 |
| MIP-1a | r=0.236<br>p=0.416 | r=0.111<br>p=0.707 | r=0.204<br>p=0.483 | r=0.313<br>p=0.276 | **r=0.257<br>p=0.020** | **r=0.357<br>p<0.001** | r=0.194<br>p=0.081 | **r=0.470<br>p<0.001** |
| IL-6 | r=0.078<br>p=0.790 | r=-0.053<br>p=0.858 | r=0.308<br>p=0.283 | r=0.192<br>p=0.512 | **r=0.354<br>p=0.001** | **r=0.243<br>p=0.028** | **r=0.277<br>p=0.012** | r=0.074<br>p=0.511 |
| TNF | r=0.194<br>p=0.507 | r=-0.026<br>p=0.929 | r=0.198<br>p=0.497 | r=0.271<br>p=0.349 | r=0.055<br>p=0.624 | r=0.035<br>p=0.752 | r=0.197<br>p=0.076 | **r=0.259<br>p=0.019** |
| IFNg | r=0.194<br>p=0.507 | r=-0.281<br>p=0.331 | r=0.146<br>p=0.618 | r=-0.173<br>p=0.554 | **r=0.346<br>p=0.001** | r=0.194<br>p=0.081 | r=0.154<br>p=0.167 | r=0.201<br>p=0.080 |
| IL-1b | r=0.310<br>p=0.280 | r=-0.024<br>p=0.934 | r=0.209<br>p=0.474 | r=0.400<br>p=0.156 | r=0.098<br>p=0.379 | r=0.099<br>p=0.375 | **r=0.230<br>p=0.037** | r=0.205<br>p=0.065 |
| IL-23 | r=0.129<br>p=0.661 | r=0.229<br>p=0.431 | r=0.108<br>p=0.714 | r=0.488<br>p=0.076 | r=0.153<br>p=0.169 | r=0.147<br>p=0.189 | r=0.158<br>p=0.158 | **r=0.208<br>p=0.005** |
| IL-12 | r=0.006<br>p=0.985 | r=0.159<br>p=0.588 | r=0.044<br>p=0.881 | **r=0.620<br>p=0.018** | r=0.030<br>p=0.790 | r=0.137<br>p=0.220 | r=0.121<br>p=0.281 | **r=0.318<br>p=0.004** |
| IL-8 | r=0.184<br>p=0.529 | r=0.183<br>p=0.531 | r=0.361<br>p=0.205 | r=0.060<br>p=0.839 | **r=0.334<br>p=0.002** | **r=0.472<br>p=0.001** | **r=0.410<br>p=0.001** | r=0.075<br>p=0.503 |

**Supplementary Table 7: IgG anti-HDL as predictor of atherosclerosis plaque burden in RA.** The role of IgG anti-HDL levels as predictor of atherosclerosis plaque burden in early RA patients was analysed by univariate and multivariate negative binomial regression models. The number of atherosclerosis plaques was entered as the dependent variable.

| | **RR** | **95% CI** | **p-value** |
|---|---|---|---|
| ***Univariate*** | | | |
| IgG anti-HDL, per unit | 1.001 | 1.000 – 1.001 | 0.023 |
| ***Multivariate*** | | | |
| IgG anti-HDL, per unit | 1.001 | 1.000 – 1.001 | 0.020 |
| Sex, women | 0.943 | 0.509 – 1.748 | 0.852 |
| Age, per year | 1.019 | 0.992 – 1.048 | 0.171 |
| Dislipemia, yes | 0.742 | 0.435 – 1.266 | 0.274 |
| Diabetes, yes | 0.001 | 0.000 – 0.001 | 0.999 |
| Hypertension, yes | 1.267 | 0.548 – 2.930 | 0.581 |
| Smoking, yes | 1.647 | 0.370 – 2.367. | 0.125 |

| | | | |
|---|---|---|---|
| Obesity, yes | 1.341 | 0.727 – 2.471 | 0.348 |

**Supplementary Table 8: Associations between antibodies against HDL and serum proteome features.** Associations between levels of antibodies against HDL or ApoA1 and levels of serum proteome features were analysed by Spearman's rank tests in RA patients. Coefficients (r) and p-values (univariate analyses, first column) and FDR-corrected (FDR=0.1) p-values (Benjamini-Hochberg, second column) are shown. Those reaching statistical significance in the latter are highlighted in bold.

| | **Anti-HDL IgG** | | **Anti-HDL IgM** | | **Anti-ApoA1 IgG** | | **Anti-ApoA1 IgM** | |
|---|---|---|---|---|---|---|---|---|
| | ***r, p univ*** | ***p BH*** | ***r, p univ*** | ***p BH*** | ***r, p univ*** | ***p BH*** | ***r, p univ*** | ***p BH*** |
| BMP6 | r=0.176<br>p=0.148 | p=0.328 | r=-0.203<br>p=0.094 | p=0.305 | r=-0.066<br>p=0.589 | p=0.949 | r=-0.221<br>p=0.068 | p=0.687 |
| ANGPT1 | r=-0.293<br>p=0.010 | **p=0.053** | r=-0.282<br>p=0.019 | p=0.130 | r=-0.248<br>p=0.040 | p=0.279 | r=-0.186<br>p=0.126 | p=0.764 |
| ADM | r=0.213<br>p=0.079 | p=0.217 | r=0.033<br>p=0.789 | p=0.908 | r=-0.104<br>p=0.397 | p=0.903 | r=-0.045<br>p=0.716 | p=0.968 |
| CD40L | r=0.094<br>p=0.440 | p=0.563 | r=-0.157<br>p=0.197 | p=0.481 | r=0.004<br>p=0.972 | p=0.993 | r=-0.005<br>p=0.967 | p=0.988 |
| SLAMF7 | r=0.393<br>p<0.01 | **p=0.053** | r=0.239<br>p=0.048 | p=0.218 | r=0.329<br>p=0.006 | p=0.085 | r=-0.045<br>p=0.713 | p=0.968 |
| PGF | r=0.330<br>p=0.006 | **p=0.049** | r=0.079<br>p=0.517 | p=0.758 | r=0.115<br>p=0.345 | p=0.835 | r=-0.042<br>p=0.732 | p=0.968 |
| ADAMTS13 | r=0.245<br>p=0.040 | **p=0.095** | r=-0.298<br>p=0.013 | p=0.127 | r=-0.236<br>p=0.051 | p=0.290 | r=-0.202<br>p=0.096 | p=0.764 |
| BOC | r=-0.032<br>p=0.797 | p=0.877 | r=-0.289<br>p=0.013 | p=0.127 | r=0.032<br>p=0.794 | p=0.993 | r=-0.193<br>p=0.113 | p=0.764 |
| IL-4RA | r=0.296<br>p=0.013 | **p=0.059** | r=0.125<br>p=0.305 | p=0.575 | r=-0.010<br>p=0.934 | p=0.993 | r=0.102<br>p=0.402 | p=0.905 |

| | | | | | | | | |
|---|---|---|---|---|---|---|---|---|
| SRC | r=0.072<br>p=0.558 | p=0.686 | r=-0.001<br>p=0.995 | p=0.995 | r=-0.097<br>p=0.426 | p=0.923 | r=-0.069<br>p=0.575 | p=0.943 |
| IL-1RA | r=0.314<br>p=0.009 | **p=0.053** | r=0.302<br>p=0.012 | p=0.127 | r=0.032<br>p=0.791 | p=0.993 | r=0.021<br>p=0.864 | p=0.968 |
| IL-6 | r=0.346<br>p=0.004 | **p=0.045** | r=0.263<br>p=0.029 | p=0.175 | r=0.270<br>p=0.025 | p=0.245 | r=0.025<br>p=0.840 | p=0.968 |
| TNFRSF10A | r=0.243<br>p=0.044 | p=0.154 | r=-0.111<br>p=0.363 | p=0.616 | r=-0.017<br>p=0.892 | p=0.993 | r=0.024<br>p=0.845 | p=0.968 |
| STK4 | r=0.126<br>p=0.303 | p=0.437 | r=0.041<br>p=0.739 | p=0.896 | r=-0.114<br>p=0.349 | p=0.835 | r=-0.066<br>p=0.589 | p=0.943 |
| IDUA | r=0.064<br>p=0.599 | p=0.698 | r=-0.216<br>p=0.075 | p=0.295 | r=-0.116<br>p=0.344 | p=0.835 | r=-0.015<br>p=0.904 | p=0.968 |
| TNFRSF11A | r=0.336<br>p=0.005 | **p=0.045** | r=0.045<br>p=0.714 | p=0.890 | r=0.083<br>p=0.500 | p=0.947 | r=-0.041<br>p=0.741 | p=0.968 |
| PAR-1 | r=0.131<br>p=0.284 | p=0.423 | r=0.006<br>p=0.958 | p=0.979 | r=-0.025<br>p=0.839 | p=0.993 | r=-0.084<br>p=0.495 | p=0.919 |
| TRAIL-R2 | r=0.180<br>p=0.138 | p=0.322 | r=0.016<br>p=0.895 | p=0.965 | r=0.022<br>p=0.856 | p=0.993 | r=0.064<br>p=0.604 | p=0.943 |
| PRSS27 | r=0.077<br>p=0.530 | p=0.660 | r=-0.242<br>p=0.046 | p=0.218 | r=-0.084<br>p=0.492 | p=0.947 | r=0.036<br>p=0.769 | p=0.968 |
| TIE2 | r=0.015<br>p=0.901 | p=0.927 | r=-0.123<br>p=0.316 | p=0.575 | r=0.063<br>p=0.609 | p=0.955 | r=-0.240<br>p=0.047 | p=0.568 |
| TF | r=-0.158<br>p=0.195 | p=0.394 | r=-0.223<br>p=0.065 | p=0.281 | r=0.003<br>p=0.981 | p=0.993 | r=-0.096<br>p=0.434 | p=0.905 |
| IL1RL2 | r=0.178<br>p=0.143 | p=0.325 | r=-0.243<br>p=0.045 | p=0.218 | r=-0.021<br>p=0.861 | p=0.993 | r=0.022<br>p=0.854 | p=0.968 |
| PDGF-b | r=-0.131<br>p=0.283 | p=0.423 | r=-0.123<br>p=0.315 | p=0.575 | r=-0.270<br>p=0.025 | p=0.245 | r=-0.220<br>p=0.049 | p=0.568 |
| IL-27 | r=0.150<br>p=0.218 | p=0.414 | r=-0.020<br>p=0.868 | p=0.955 | r=0.066<br>p=0.587 | p=0.949 | r=-0.187<br>p=0.124 | p=0.764 |

| | | | | | | | | |
|---|---|---|---|---|---|---|---|---|
| IL-17D | r=0.068<br>p=0.576 | p=0.698 | r=-0.065<br>p=0.594 | p=0.819 | r=-0.023<br>p=0.852 | p=0.993 | r=0.024<br>p=0.847 | p=0.968 |
| CXCL1 | r=0.333<br>p=0.005 | **p=0.045** | r=-0.042<br>p=0.732 | p=0.896 | r=0.149<br>p=0.221 | p=0.835 | r=0.125<br>p=0.307 | p=0.828 |
| Gal-9 | r=0.306<br>p=0.010 | **p=0.053** | r=0.124<br>p=0.311 | p=0.575 | r=0.121<br>p=0.321 | p=0.835 | r=-0.065<br>p=0.596 | p=0.943 |
| GIF | r=0.283<br>p=0.018 | **p=0.078** | r=-0.199<br>p=0.102 | p=0.045 | r=-0.142<br>p=0.244 | p=0.835 | r=-0.350<br>p=0.003 | p=0.273 |
| SCF | r=-0.458<br>p=0.001 | **p=0.030** | r=-0.414<br>p<0.001 | **p=0.032** | r=-0.296<br>p=0.014 | p=0.245 | r=-0.160<br>p=0.189 | p=0.775 |
| IL-18 | r=0.343<br>p=0.004 | **p=0.045** | r=0.288<br>p=0.017 | p=0.130 | r=0.273<br>p=0.023 | p=0.245 | r=0.057<br>p=0.642 | p=0.957 |
| FGF21 | r=0.255<br>p=0.035 | **p=0.094** | r=0.177<br>p=0.145 | p=0.402 | r=-0.028<br>p=0.818 | p=0.993 | r=-0.076<br>p=0.535 | p=0.942 |
| PIgR | r=-0.021<br>p=0.866 | p=0.916 | r=-0.156<br>p=0.201 | p=0.481 | r=-0.076<br>p=0.534 | p=0.949 | r=-0.011<br>p=0.926 | p=0.968 |
| RAGE | r=0.161<br>p=0.187 | p=0.394 | r=-0.161<br>p=0.186 | p=0.481 | r=-0.020<br>p=0.870 | p=0.993 | r=0.119<br>p=0.332 | p=0.828 |
| SOD2 | r=0.147<br>p=0.227 | p=0.414 | r=-0.171<br>p=0.159 | p=0.425 | r=0.174<br>p=0.153 | p=0.696 | r=0.092<br>p=0.451 | p=0.912 |
| CTRC | r=0.122<br>p=0.320 | p=0.455 | r=0.008<br>p=0.950 | p=0.979 | r=0.065<br>p=0.595 | p=0.949 | r=0.031<br>p=0.800 | p=0.968 |
| FGF23 | r=0.126<br>p=0.302 | p=0.437 | r=-0.11<br>p=0.366 | p=0.616 | r=-0.072<br>p=0.556 | p=0.949 | r=0.009<br>p=0.941 | p=0.973 |
| SPON2 | r=0.312<br>p=0.008 | **p=0.053** | r=-0.096<br>p=0.431 | p=0.676 | r=0.003<br>p=0.980 | p=0.993 | r=-0.073<br>p=0.549 | p=0.942 |
| GH | r=0.103<br>p=0.398 | p=0.532 | r=0.003<br>p=0.982 | p=0.992 | r=0.026<br>p=0.833 | p=0.993 | r=-0.016<br>p=0.893 | p=0.968 |
| FS | r=0.153<br>p=0.210 | p=0.414 | r=-0.193<br>p=0.112 | p=0.339 | r=-0.042<br>p=0.734 | p=0.993 | r=-0.163<br>p=0.182 | p=0.775 |

| | | | | | | | | |
|---|---|---|---|---|---|---|---|---|
| GLO1 | r=0.029<br>p=0.813 | p=0.877 | r=-0.214<br>p=0.078 | p=0.295 | r=0.048<br>p=0.695 | p=0.993 | r=-0.155<br>p=0.203 | p=0.775 |
| CD84 | r=-0.016<br>p=0.894 | p=0.927 | r=-0.177<br>p=0.145 | p=0.402 | r=-0.126<br>p=0.302 | p=0.835 | r=-0.145<br>p=0.236 | p=0.775 |
| PAPPA | r=0.142<br>p=0.245 | p=0.414 | r=-0.113<br>p=0.357 | p=0.616 | r=-0.066<br>p=0.589 | p=0.949 | r=-0.098<br>p=0.421 | p=0.905 |
| SERPINA12 | r=0.014<br>p=0.910 | p=0.927 | r=-0.068<br>p=0.581 | p=0.813 | r=0.052<br>p=0.674 | p=0.993 | r=-0.085<br>p=0.489 | p=0.919 |
| REN | r=0.184<br>p=0.129 | p=0.317 | r=-0.279<br>p=0.020 | p=0.130 | r=-0.082<br>p=0.500 | p=0.947 | r=-0.180<br>p=0.138 | p=0.775 |
| DECR1 | r=0.181<br>p=0.137 | p=0.322 | r=0.048<br>p=0.696 | p=0.879 | r=0.048<br>p=0.693 | p=0.993 | r=0.192<br>p=0.114 | p=0.764 |
| MERTK | r=0.143<br>p=0.241 | p=0.414 | r=-0.088<br>p=0.473 | p=0.729 | r=0.104<br>p=0.397 | p=0.903 | r=-0.193<br>p=0.112 | p=0.764 |
| KIM1 | r=0.323<br>p=0.007 | **p=0.053** | r=-0.102<br>p=0.406 | p=0.651 | r=0.012<br>p=0.920 | p=0.993 | r=-0.151<br>p=0.215 | p=0.775 |
| THBS2 | r=0.465<br>p<0.001 | **p=0.030** | r=0.139<br>p=0.254 | p=0.550 | r=0.002<br>p=0.990 | p=0.993 | r=-0.117<br>p=0.337 | p=0.828 |
| TM | r=0.244<br>p=0.043 | p=0.154 | r=0.055<br>p=0.625 | p=0.825 | r=-0.013<br>p=0.913 | p=0.993 | r=-0.062<br>p=0.612 | p=0.943 |
| VSIG2 | r=0.166<br>p=0.172 | p=0.372 | r=-0.026<br>p=0.835 | p=0.938 | r=-0.092<br>p=0.454 | p=0.947 | r=-0.151<br>p=0.215 | p=0.775 |
| AMBP | r=0.131<br>p=0.284 | p=0.423 | r=-0.159<br>p=0.192 | p=0.481 | r=-0.119<br>p=0.332 | p=0.835 | r=-0.155<br>p=0.205 | p=0.775 |
| PRELP | r=0.013<br>p=0.917 | p=0.927 | r=-0.293<br>p=0.014 | p=0.127 | r=-0.135<br>p=0.269 | p=0.835 | r=-0.082<br>p=0.505 | p=0.919 |
| HO-1 | r=0.081<br>p=0.799 | p=0.877 | r=-0.062<br>p=0.612 | p=0.825 | r=0.304<br>p=0.011 | p=0.245 | r=-0.167<br>p=0.169 | p=0.775 |
| XCL1 | r=0.237<br>p=0.050 | p=0.156 | r=0.131<br>p=0.283 | p=0.574 | r=0.117<br>p=0.336 | p=0.835 | r=0.000<br>p=0.999 | p=0.999 |

| | | | | | | | | |
|---|---|---|---|---|---|---|---|---|
| IL-16 | r=0.141<br>p=0.246 | p=0.414 | r=0.133<br>p=0.277 | p=0.574 | r=0.265<br>p=0.027 | p=0.245 | r=0.325<br>p=0.007 | p=0.273 |
| SORT1 | r=0.004<br>p=0.977 | p=0.977 | r=-0.068<br>p=0.581 | p=0.813 | r=-0.144<br>p=0.237 | p=0.835 | r=-0.118<br>p=0.336 | p=0.828 |
| CEACAM8 | r=0.218<br>p=0.072 | p=0.204 | r=0.149<br>p=0.220 | p=0.500 | r=0.001<br>p=0.993 | p=0.993 | r=0.057<br>p=0.641 | p=0.957 |
| PTX3 | r=0.274<br>p=0.061 | p=0.185 | r=0.249<br>p=0.039 | p=0.208 | r=-0.067<br>p=0.582 | p=0.949 | r=0.070<br>p=0.570 | p=0.943 |
| PSGL-1 | r=0.138<br>p=0.257 | p=0.423 | r=-0.204<br>p=0.094 | p=0.305 | r=0.129<br>p=0.289 | p=0.835 | r=-0.038<br>p=0.759 | p=0.968 |
| CCL17 | r=0.100<br>p=0.412 | p=0.543 | r=-0.059<br>p=0.629 | p=0.825 | r=0.163<br>p=0.181 | p=0.784 | r=0.314<br>p=0.009 | p=0.273 |
| CCL3 | r=0.146<br>p=0.231 | p=0.414 | r=0.367<br>p=0.002 | **p=0.060** | r=0.054<br>p=0.658 | p=0.993 | r=-0.086<br>p=0.484 | p=0.919 |
| MMP7 | r=0.207<br>p=0.088 | p=0.235 | r=-0.125<br>p=0.305 | p=0.575 | r=0.126<br>p=0.301 | p=0.835 | r=-0.014<br>p=0.908 | p=0.968 |
| IgG FgR IIb | r=0.090<br>p=0.460 | p=0.581 | r=-0.072<br>p=0.552 | p=0.797 | r=0.034<br>p=0.780 | p=0.993 | r=0.016<br>p=0.893 | p=0.968 |
| ITGB1BP2 | r=0.297<br>p=0.013 | **p=0.059** | r=0.217<br>p=0.074 | p=0.295 | r=0.244<br>p=0.044 | p=0.279 | r=0.025<br>p=0.841 | p=0.968 |
| DCN | r=0.222<br>p=0.066 | p=0.193 | r=-0.145<br>p=0.234 | p=0.519 | r=0.007<br>p=0.955 | p=0.993 | r=-0.127<br>p=0.300 | p=0.828 |
| Dkk-1 | r=0.065<br>p=0.597 | p=0.698 | r=-0.206<br>p=0.089 | p=0.305 | r=-0.019<br>p=0.880 | p=0.993 | r=-0.143<br>p=0.242 | p=0.775 |
| LPL | r=-0.290<br>p=0.012 | **p=0.049** | r=-0.294<br>p=0.013 | p=0.127 | r=-0.284<br>p=0.018 | p=0.245 | r=-0.084<br>p=0.493 | p=0.919 |
| PRSS8 | r=0.054<br>p=0.661 | p=0.761 | r=-0.102<br>p=0.405 | p=0.651 | r=-0.252<br>p=0.037 | p=0.279 | r=-0.144<br>p=0.239 | p=0.775 |
| AGRP | r=0.230<br>p=0.040 | p=0.151 | r=0.152<br>p=0.211 | p=0.492 | r=0.128<br>p=0.296 | p=0.835 | r=0.046<br>p=0.912 | p=0.968 |

| | | | | | | | | |
|---|---|---|---|---|---|---|---|---|
| HB-EGF | r=0.137<br>p=0.262 | p=0.423 | r=-0.282<br>p=0.019 | p=0.130 | r=-0.083<br>p=0.496 | p=0.947 | r=-0.118<br>p=0.333 | p=0.828 |
| GDF-2 | r=-0.029<br>p=0.810 | p=0.877 | r=-0.177<br>p=0.146 | p=0.402 | r=-0.002<br>p=0.987 | p=0.993 | r=-0.075<br>p=0.543 | p=0.942 |
| FABP2 | r=-0.111<br>p=0.364 | p=0.494 | r=-0.084<br>p=0.494 | p=0.739 | r=0.013<br>p=0.915 | p=0.993 | r=-0.141<br>p=0.247 | p=0.775 |
| THPO | r=-0.202<br>p=0.095 | p=0.247 | r=-0.393<br>p<0.001 | **p=0.045** | r=-0.088<br>p=0.470 | p=0.947 | r=-0.124<br>p=0.309 | p=0.828 |
| MARCO | r=0.145<br>p=0.236 | p=0.414 | r=-0.031<br>p=0.801 | p=0.911 | r=0.217<br>p=0.074 | p=0.396 | r=-0.049<br>p=0.691 | p=0.968 |
| GT | r=0.159<br>p=0.191 | p=0.394 | r=0.015<br>p=0.902 | p=0.965 | r=0.046<br>p=0.705 | p=0.993 | r=0.163<br>p=0.181 | p=0.775 |
| BNP | r=0.135<br>p=0.270 | p=0.423 | r=0.118<br>p=0.336 | p=0.599 | r=-0.138<br>p=0.259 | p=0.835 | r=0.095<br>p=0.438 | p=0.905 |
| MMP12 | r=0.352<br>p=0.003 | **p=0.045** | r=0.346<br>p=0.004 | **p=0.072** | r=0.328<br>p=0.006 | p=0.245 | r=0.237<br>p=0.050 | p=0.568 |
| ACE2 | r=0.047<br>p=0.704 | p=0.800 | r=-0.007<br>p=0.955 | p=0.979 | r=-0.117<br>p=0.340 | p=0.835 | r=-0.050<br>p=0.681 | p=0.968 |
| PD-L2 | r=0.148<br>p=0.226 | p=0.414 | r=-0.035<br>p=0.777 | p=0.906 | r=0.072<br>p=0.559 | p=0.949 | r=-0.001<br>p=0.991 | p=0.999 |
| CSTL1 | r=0.232<br>p=0.050 | p=0.156 | r=-0.038<br>p=0.777 | p=0.906 | r=0.099<br>p=0.418 | p=0.923 | r=0.010<br>p=0.990 | p=0.968 |
| hOSCAR | r=0.361<br>p=0.002 | **p=0.036** | r=0.250<br>p=0.038 | p=0.208 | r=0.241<br>p=0.046 | p=0.279 | r=0.236<br>p=0.033 | p=0.568 |
| TNFRSF13B | r=0.362<br>p=0.002 | **p=0.036** | r=0.039<br>p=0.750 | p=0.898 | r=0.069<br>p=0.576 | p=0.949 | r=-0.051<br>p=0.677 | p=0.968 |
| TGM2 | r=0.028<br>p=0.820 | p=0.877 | r=-0.208<br>p=0.086 | p=0.305 | r=-0.042<br>p=0.734 | p=0.993 | r=-0.273<br>p=0.023 | p=0.523 |
| LEP | r=-0.117<br>p=0.339 | p=0.474 | r=-0.131<br>p=0.284 | p=0.574 | r=-0.269<br>p=0.020 | p=0.245 | r=-0.142<br>p=0.244 | p=0.775 |

| | | | | | | | | |
|---|---|---|---|---|---|---|---|---|
| CA5A | r=0.098 p=0.424 | p=0.551 | r=-0.053 p=0.664 | p=0.851 | r=-0.185 p=0.127 | p=0.608 | r=-0.138 p=0.257 | p=0.779 |
| HSP27 | r=0.186 p=0.127 | p=0.317 | r=0.058 p=0.635 | p=0.825 | r=0.020 p=0.867 | p=0.993 | r=-0.037 p=0.761 | p=0.968 |
| CD4 | r=0.343 p=0.001 | **p=0.030** | r=0.083 p=0.496 | p=0.739 | r=0.246 p=0.042 | p=0.279 | r=0.012 p=0.924 | p=0.968 |
| NEMO | r=0.131 p=0.284 | p=0.423 | r=-0.020 p=0.871 | p=0.955 | r=-0.033 p=0.787 | p=0.993 | r=0.103 p=0.399 | p=0.905 |
| VEGFD | r=0.115 p=0.348 | p=0.479 | r=-0.351 p=0.003 | **p=0.068** | r=-0.204 p=0.092 | p=0.465 | r=-0.110 p=0.368 | p=0.881 |
| PARP-1 | r=0.237 p=0.050 | p=0.156 | r=0.011 p=0.930 | p=0.979 | r=0.010 p=0.935 | p=0.993 | r=0.100 p=0.415 | p=0.905 |
| HAOX1 | r=0.067 p=0.583 | p=0.698 | r=0.101 p=0.408 | p=0.651 | r=-0.126 p=0.304 | p=0.835 | r=0.033 p=0.788 | p=0.968 |

**Supplementary Table 9: List of proteins associated with IgG or IgM anti-HDL antibodies in RA.** Serum proteins found to be associated with IgG (n=23) or IgM (n=5) anti-HDL antibodies are listed along with their KEGG ortholog codes and most representative functions.

| | **Protein** | **KEGG ortholog** | **Functions** |
|---|---|---|---|
| **Anti-HDL IgG** | SCF | K05461 | Cell survival and proliferation, hematopoiesis, stem cell maintenance, gametogenesis, mast cell development, migration and function |
| | THBS2 | K04659 | Mediates cell-to-cell and cell-to-matrix interactions. Ligand for CD36 mediating antiangiogenic properties |
| | CD4 | K06454 | Immune activation |
| | hOSCAR | K14377 | Osteoclastogenesis, remodelling |
| | TNFRSF13B | K05150 | Stimulation of B- and T-cell function and the regulation of humoral immunity |
| | MMP12 | K01413 | Extracellular matrix remodelling |
| | IL-6 | K05405 | Immune activation |
| | IL-18 | K10030 | Immune activation |
| | TNFRSF11A | K05147 | Osteoclastogenesis, remodelling, interactions between T-cells and dendritic cells |
| | CXCL1 | K05505 | Neutrophil activation and migration; endothelial activation |
| | PGF | K16859 | Angiogenesis and endothelial cell growth, stimulating their proliferation and migration |
| | KIM1 | K20413 | T-helper cell development and the regulation of asthma and allergic diseases<br>Kidney injury and repair |
| | SPON2 | K24428 | Cell adhesion<br>Opsonization<br>Innate immune response, LPS-R<br>Extracellular matrix homeostasis |

| | | | |
|---|---|---|---|
| | IL-1RA | K05071 | Immune regulation |
| | ANGPT1 | K05465 | Regulation of angiogenesis, endothelial cell survival, proliferation, migration, adhesion and cell spreading, reorganization of the actin cytoskeleton, but also maintenance of vascular quiescence<br>Vascular homeostasis |
| | Gal-9 | K10093 | Inhibition angiogenesis<br>Th1/Th2 polarization |
| | SLAMF7 | K06733 | Regulation and interconnection of both innate and adaptive immune response |
| | LPL | K01059 | Triglyceride and TRL metabolism<br>Lipid clearance |
| | IL-4RA | K05071 | Immune regulation |
| | ITGB1BP2 | K16735 | Maturation and differentiation of muscle cells |
| | CBLIF | K14615 | Vitamin digestion and absorption |
| | FGF21 | K22429 | Glucose metabolism<br>Adipose tissue homeostasis |
| | ADAMTS13 | K08627 | vWF production<br>Extracellular matrix homeostasis |
| **Anti-HDL IgM** | SCF | K05461 | Cell survival and proliferation, hematopoiesis, stem cell maintenance, gametogenesis, mast cell development, migration and function |
| | THPO | K06854 | Proliferation and maturation of megakaryocytes from their committed progenitor cells.<br>Major physiological regulator of circulating platelets |
| | CCL3 | K05408 | Monocyte chemotaxis |
| | VEGFD | K05449 | Angiogenesis, lymphangiogenesis and endothelial cell growth, stimulating their proliferation and migration, and also has effects on the permeability of blood vessels<br>Vascular homeostasis |
| | MMP12 | K01413 | Extracellular matrix remodeling |

**Supplementary Table 10: List of candidate transcription factors for the proteins analyzed.** Transcription factors identified to be common for the proteins analyzed using the TRRUST database.

| **Abbreviation** | **Transcription factor** | **# overlapped genes** | **p-value** | **FDR** |
|---|---|---|---|---|
| RELA | v-rel reticuloendotheliosis viral oncogene homolog A (avian) | 6 | $9.41 \cdot 10^{-7}$ | $8.47 \cdot 10^{-6}$ |
| NFKB1 | nuclear factor of kappa light polypeptide gene enhancer in B-cells 1 | 5 | $2.17 \cdot 10^{-5}$ | $9.77 \cdot 10^{-5}$ |
| STAT3 | signal transducer and activator of transcription 3 (acute-phase response factor) | 3 | 0.0005 | 0.0016 |
| ATF4 | activating transcription factor 4 (tax-responsive enhancer element B67) | 2 | 0.0007 | 0.0016 |
| PPARA | peroxisome proliferator-activated receptor alpha | 2 | 0.0009 | 0.0016 |
| SPI1 | spleen focus forming virus (SFFV) proviral integration oncogene spi1 | 2 | 0.0023 | 0.0035 |
| CREB1 | cAMP responsive element binding protein 1 | 2 | 0.0048 | 0.0062 |
| JUN | jun proto-oncogene | 2 | 0.0129 | 0.0145 |
| SP1 | Sp1 transcription factor | 3 | 0.0168 | 0.0168 |

### SUPPLEMENTARY FIGURE LEGENDS

**Supplementary Figure 1: Levels of antibodies against HDL particles in clinical RA.** Levels of IgG anti-HDL and anti-ApoA1 (both IgG and IgM) in early RA (RA) and established, long-lasting RA (LRA) are shown. Bars represent 25th percentile (lower), median and 75th percentile (upper). Differences were assessed by Mann-Withney U tests.

**Supplementary Figure 2: PON1 activity across study groups.** Levels PON1 enzymatic activity (U) in HC, CSA individuals and early RA patients are shown. Bars represent 25$^{th}$ percentile (lower), median and 75$^{th}$ percentile (upper). Differences were assessed by Kruskal-Wallis tests with Dunn-Bonferroni post-hoc tests. The p-values from the latter were indicated as follows: * $p<0.050$, and *** $p<0.001$.

**Supplementary Figure 3: Network analyses of proteomic signatures and atherosclerosis status.** The interactions among proteins found to be associated with IgG anti-HDL levels were depicted in networks for patients without atherosclerosis (n=31) and with atherosclerosis (n=46). Each node corresponds to a protein and the lines between nodes illustrate the strength (width) and type (red: positive, blue: negative) of the correlations between each pair of nodes. The relative position of the nodes parallels its degree of correlation, that is, nodes more closely correlated locate closer to each other.
